# Safety and immunogenicity of the protein-based PHH-1V compared to BNT162b2 as a heterologous SARS-CoV-2 booster vaccine in adults vaccinated against COVID-19: a multicentre, randomised, double-blind, non-inferiority phase IIb trial

**DOI:** 10.1101/2022.07.05.22277210

**Authors:** Júlia Corominas, Carme Garriga, Antoni Prenafeta, Alexandra Moros, Manuel Cañete, Antonio Barreiro, Luis González-González, Laia Madrenas, Irina Güell, Bonaventura Clotet, Nuria Izquierdo-Useros, Dàlia Raïch-Regué, Marçal Gallemí, Julià Blanco, Edwards Pradenas, Benjamin Trinité, Julia G Prado, Oscar Blanch-Lombarte, Raúl Pérez-Caballero, Montserrat Plana, Ignasi Esteban, Carmen Pastor-Quiñones, Xavier Núñez-Costa, Rachel Abu Taleb, Paula McSkimming, Alex Soriano, Jocelyn Nava, Jesse Omar Anagua, Rafel Ramos, Ruth Martí Lluch, Aida Corpes Comes, Susana Otero Romero, Xavier Martinez Gomez, Carla Sans-Pola, José Moltó, Susana Benet, Lucía Bailón, Jose R Arribas, Alberto M Borobia, Javier Queiruga Parada, Jorge Navarro-Pérez, Maria José Forner Giner, Rafael Ortí Lucas, María del Mar Vázquez Jiménez, Salvador Oña Compán, Melchor Alvarez-Mon, Daniel Troncoso, Eunate Arana-Arri, Susana Meijide, Natale Imaz-Ayo, Patricia Muñoz García, Sofía de la Villa Martínez, Sara Rodríguez Fernández, Teresa Prat, Èlia Torroella, Laura Ferrer

## Abstract

**Background:** A SARS-CoV-2 protein-based heterodimer vaccine, PHH-1V, has been shown to be safe and well-tolerated in healthy young adults in a first-in-human, Phase I/IIa study dose-escalation trial. Here, we report the interim results of the Phase IIb HH-2, where the immunogenicity and safety of a heterologous booster with PHH-1V is assessed versus a homologous booster with BNT162b2 at 14, 28 and 98 days after vaccine administration.

**Methods:** The HH-2 study is an ongoing multicentre, randomised, active-controlled, double-blind, non-inferiority Phase IIb trial, where participants 18 years or older who had received two doses of BNT162b2 were randomly assigned in a 2:1 ratio to receive a booster dose of vaccine —either heterologous (PHH-1V group) or homologous (BNT162b2 group)— in 10 centres in Spain. Eligible subjects were allocated to treatment stratified by age group (18-64 versus ≥65 years) with approximately 10% of the sample enrolled in the older age group. The primary endpoints were humoral immunogenicity measured by changes in levels of neutralizing antibodies (PBNA) against the ancestral Wuhan-Hu-1 strain after the PHH-1V or the BNT162b2 boost, and the safety and tolerability of PHH-1V as a boost. The secondary endpoints were to compare changes in levels of neutralizing antibodies against different variants of SARS-CoV-2 and the T-cell responses towards the SARS-CoV-2 spike glycoprotein peptides. The exploratory endpoint was to assess the number of subjects with SARS-CoV-2 infections ≥14 days after PHH-1V booster. This study is ongoing and is registered with ClinicalTrials.gov, NCT05142553.

**Findings:** From 15 November 2021, 782 adults were randomly assigned to PHH-1V (n=522) or BNT162b2 (n=260) boost vaccine groups. The geometric mean titre (GMT) ratio of neutralizing antibodies on days 14, 28 and 98, shown as BNT162b2 active control versus PHH-1V, was, respectively, 1·68 (p<0·0001), 1·31 (p=0·0007) and 0·86 (p=0·40) for the ancestral Wuhan-Hu-1 strain; 0·62 (p<0·0001), 0·65 (p<0·0001) and 0·56 (p=0·003) for the Beta variant; 1·01 (p=0·92), 0·88 (p=0·11) and 0·52 (p=0·0003) for the Delta variant; and 0·59 (p=<0·0001), 0·66 (p<0·0001) and 0·57 (p=0·0028) for the Omicron BA.1 variant. Additionally, PHH-1V as a booster dose induced a significant increase of CD4^+^ and CD8^+^ T-cells expressing IFN-γ on day 14. There were 458 participants who experienced at least one adverse event (89·3%) in the PHH-1V and 238 (94·4%) in the BNT162b2 group. The most frequent adverse events were injection site pain (79·7% and 89·3%), fatigue (27·5% and 42·1%) and headache (31·2 and 40·1%) for the PHH-1V and the BNT162b2 groups, respectively. A total of 52 COVID-19 cases occurred from day 14 post-vaccination (10·14%) for the PHH-1V group and 30 (11·90%) for the BNT162b2 group (p=0·45), and none of the subjects developed severe COVID-19.

**Interpretation:** Our interim results from the Phase IIb HH-2 trial show that PHH-1V as a heterologous booster vaccine, when compared to BNT162b2, although it does not reach a non-inferior neutralizing antibody response against the Wuhan-Hu-1 strain at days 14 and 28 after vaccination, it does so at day 98. PHH-1V as a heterologous booster elicits a superior neutralizing antibody response against the previous circulating Beta and the currently circulating Omicron BA.1 SARS-CoV-2 variants in all time points assessed, and for the Delta variant on day 98 as well. Moreover, the PHH-1V boost also induces a strong and balanced T-cell response. Concerning the safety profile, subjects in the PHH-1V group report significantly fewer adverse events than those in the BNT162b2 group, most of mild intensity, and both vaccine groups present comparable COVID-19 breakthrough cases, none of them severe.

**Funding:** HIPRA SCIENTIFIC, S.L.U.

## Introduction

The severe acute respiratory syndrome coronavirus 2 (SARS-CoV-2) identified in Wuhan, China, in December 2019, has brought an urgency to prophylactic measures globally. Consequently, the world has witnessed a striking change in the speed of vaccine development targeting the SARS-CoV-2’s membrane spike protein, particularly its receptor binding domain (RBD), which interacts with the angiotensin converting enzyme 2 (ACE2) receptor in the host cells and generates a neutralizing antibody response.^1,2^ There is a positive correlation between the levels of SARS-CoV-2 neutralizing antibodies induced by vaccination and the conferred level of protection, and antibody levels have been previously used as endpoints for many viral vaccines.^3^

Clinical trials for several approved SARS-CoV-2 vaccines also revealed the induction of a rapid and specific T-cell-mediated response in most participants noted from day 14 post-vaccination, asserting its value as a basis for immunological memory and its role in the rapid response to pathogen re-exposure, conferring protection from severe disease even with lower levels of antibodies.^4-7^

The types of vaccines developed for SARS-CoV-2 carry the virus’s genetic material, contain inactivated virus or are protein-based. The first type contains the genetic information for the biosynthesis of the spike glycoprotein, such as mRNA (BNT162b2 [Pfizer/BioNTech] and mRNA-1273 [Moderna]) and adenoviral vector vaccines (Ad26.COV2.S [Janssen] and ChAdOx [Oxford-AstraZeneca]). ^8^ Vaccines based on the mRNA technology have inherent limitations: the liability of the mRNA molecule requires that mRNA-based vaccines are encapsulated in lipid nanoparticles and kept at very low temperatures (between -80 ºC and -20 ºC).^9^ Regarding the inactivated virus vaccines, the BBIBP-CorV (Sinopharm), Coronavac (Sinovac) and Covaxin (Bharat Biotech) have been approved by the World Health Organization (WHO) for emergency use. On the other hand, the protein-based vaccines contain the spike protein in different forms and combinations with adjuvants. The NVX-CoV2373 (Novavax) corresponds to a subunit vaccine, containing the full-length spike protein with the Matrix M1 adjuvant, also approved by the WHO and EMA. The CoV2 preS dTM adjuvanted vaccine (Sanofi/GSK) is a prefusion spike recombinant protein vaccine recently approved by the EMA. In contrast to mRNA, protein-based vaccines can be stored under refrigerated conditions.^6,10,11^

Although various SARS-CoV-2 vaccines have been approved, there is still the utmost need for options able to meet global demands of immunizations.^12^ Vaccination campaigns protect against severe disease, reduce viral load and therefore lower transmission risk, which implies a public health benefit.^13^ Booster vaccination has become necessary because immunity diminishes with time and new variants emerge. Increasing evidence shows that heterologous vaccination is safe, with similar reactogenicity and adverse events compared to a homologous schedules.^14-17^ Several recent clinical studies indicate that heterologous schedules may, in fact, elicit a more robust immune response and higher antibody titres, with consequent higher effectiveness, supporting a mix-and-match approach.^18-20^ These data suggest that there is a strong case for more vaccine platforms to broaden vaccination options worldwide.

Recombinant protein-based vaccines possess a good safety profile, no risk of genome integration, no live components, and are suitable for people with compromised immune systems, showing high productivity yields and good stability profiles.^21,22^ PHH-1V is a protein-based vaccine intended for the prevention of COVID-19 caused by SARS-CoV-2. This vaccine is based on a fusion heterodimer consisting of the spike RBD sequence from the SARS-CoV-2 Beta (B.1.351) and Alpha (B.1.1.7) variants, formulated with an oil-in-water emulsion based on squalene (SQBA) intended for intramuscular administration.^23-25^ PHH-1V comprises three key mutations of high relevance for previously and currently circulating SARS-CoV-2 variants: K417N, E484K and N501Y.^26^ Previous studies revealed that the E484K and N501Y mutations are present in the RBD of the spike protein of Beta, Gamma, and Mu variants; and both the K417N and N501Y mutations present in the PHH-1V antigen are common to the Beta and to the currently predominant Omicron variants.^27^

A first-in-human Phase I/IIa study dose-escalation, randomized, double-blinded, active-comparator controlled clinical trial in 30 healthy adults demonstrated that the PHH-1V vaccine is safe and well-tolerated in healthy young adults, with even fewer reported solicited adverse events compared to the control, BNT162b2^28^.

The HH-2 study is an ongoing multicentre, randomised, active-controlled, double-blind, non-inferiority Phase IIb trial that aims to assess the immunogenicity and safety of PHH-1V as a heterologous boost versus a homologous boost in individuals after receiving a primary vaccination series of BNT162b2. Here we show interim data up to 98 days after administration of the booster for the Phase IIb HH-2 study.

## Methods

### Study design and participants

This multicentre, randomised, active-controlled, double-blind, non-inferiority Phase IIb trial where immunogenicity and safety of the PHH-1V vaccine were assessed, was carried out in 10 centres in Spain (appendix pp.10-11 Supplementary material).

Eligibility criteria were individuals aged 18 years or older, who had received two doses of the BNT162b2 vaccine at least 182 days and less than 365 days after their second dose; Body Mass Index (BMI) between 18 and 40 kg/m^2^; negative SARS-CoV-2 PCR test at the time of enrolment; willingness to avoid all other vaccines within 4 weeks before and after vaccination in this study (seasonal influenza vaccination was allowed if it was received at least 14 days before or after the study booster). Key exclusion criteria included pregnancy or breastfeeding; an ongoing serious psychiatric condition; history of respiratory disease requiring daily medications; history of significant cardiovascular disease; history of neurological or neurodevelopmental conditions; ongoing malignancy or recent diagnosis of malignancy in the last five years excluding basal cell and squamous cell carcinoma of the skin; any confirmed or suspected autoimmune, immunosuppressive or immunodeficiency disease/condition (iatrogenic or congenital) including human immunodeficiency virus infection; use of immunosuppressants; coagulation or bleeding disorder; chronic liver disease; history of SARS-CoV-2 infection; close contact with anyone positive for SARS-CoV-2 infection within 15 days before screening and life expectancy of less than 12 months. Full inclusion and exclusion criteria are provided in the study protocol (Supplementary data).

The trial was conducted in accordance with the Declaration of Helsinki, the Good Clinical Practice guidelines, and national regulations. The study protocol was reviewed and approved by the Spanish Agency of Medicines and Medical Devices (AEMPS) as well as Independent Ethics Committee from the Hospital Clínic de Barcelona. Written informed consent was obtained from all participants before enrolment. The biologic biosafety committee of the Research Institute Germans Trias i Pujol approved the execution of SARS-CoV-2 experiments at the BSL3 laboratory of the Centre for Bioimaging and Comparative Medicine (CSB-20-015-M8).

### Randomisation and masking

This study was double-blinded; participants, site staff, including clinical staff involved in study drug preparation and administration, laboratory analysis staff, the sponsor, and the Clinical Research Organisation (CRO) were blinded to treatment assignment/allocation. Unblinded hospital pharmacists or other qualified personnel prepared the booster dose, and unblinded site staff members, who were not otherwise involved with study procedures (except for blood extraction), administered treatment to subjects.

Subjects were randomly assigned in a 2:1 ratio to receive a booster dose of vaccine (third immunization) either with the HIPRA PHH-1V vaccine (PHH-1V group) or with the Pfizer/BioNTech BNT162b2 vaccine (BNT162b2 group). Subjects were allocated to treatment using an Interactive Response Technology (IRT). The allocation sequence was stratified by age group: approximately 90% of the total recruited participants were in the 18 to 64 years group, and 10% in the group 65 years or older. This article refers to the data analysis obtained on days 14, 28 and 98.

### Study vaccine

The PHH-1V vaccine was provided in a vial containing one dose of 0·5 ml (40 μg), ready to use, and stored at 2-8 ºC. The adjuvant is an oil-in-water emulsion based on squalene produced by HIPRA (SQBA).

Comirnaty was supplied as a frozen suspension in a multidose vial that must be thawed and diluted before use. One vial (0·45 mL) contained 6 doses of 0·3 mL after dilution. Comirnaty was stored in an ultra-low temperature freezer between -90 to -60 ºC, until the expiry date printed on the label. Alternatively, Comirnaty could be stored at -25 to -15 ºC for up to 2 weeks. Vials were kept frozen and protected from light, in the original package, until ready to use.

A label was used to mask the syringe to visually distinguish the two treatments. The blind was broken as soon as the last study participant reached day 28 after the study booster dose. Participants would then know which vaccine they had received and were able to decide whether they wanted to receive, or not, a commercial COVID 19-vaccine. In the event a participant decided to receive a commercial COVID-19 vaccine booster according to the vaccination schedule established by local authorities, an extra visit was scheduled, with the recommendation to wait 3 months after administration of HIPRA’s vaccine to receive the commercial booster.

### Procedures

The study visits were scheduled on day 0, day 14, day 28, day 98, day 182 and day 364. This article reports data up to day 98. Participants who, at that moment, met eligibility criteria were vaccinated in the visit on day 0. The BNT162b2 vaccine was given as a 0·3 mL (30 μg) and the PHH-1V as a 0·5 mL (40 μg) by intramuscular injection into the deltoid muscle. The first 30 participants were observed for 60 minutes and monitored during the following 72 hours by phone. Other participants were observed for 30 minutes and contacted again on day 7. During the day-0 visit, participants were given a hard copy diary to record local and systemic solicited reactions within the 7 days after vaccination.

The neutralization titres of antibodies were determined by the inhibitory dilution 50 (ID_50,_ reported as the half maximal inhibitory dilution, which is the reciprocal dilution of the test serum required to inhibit the virus effect by 50%) using a pseudovirion-based neutralisation assay (PBNA) as described previously.^29^ The immunogenicity against the SARS-CoV-2 spike glycoprotein was assessed by the percentage of subjects having a ≥4-fold increase in the binding antibodies titre 14, 28 and 98 days after boosting using the Elecsys Anti-SARS-CoV-2 S immunoassay (Roche Diagnostics). The titre of neutralizing antibodies was also analysed in a subset of subjects, corresponding to approximately 20% of the total subjects included in the study, representing the first 58 participants vaccinated during this HH-2 trial (for BNT162b2 active control, n=24; for PHH-1, n=34) by infectious SARS-CoV-2 neutralisation test (VNA) and measured as ID_50_. The geometric mean titre (GMT) and the geometrical mean fold rise (GMFR) were calculated for each parameter.

The safety assessment included the incidence of solicited local and systemic reactions, unsolicited local and systemic adverse events reported in the patient’s diary. Additionally, the incidence of SARS-CoV-2 infections, including severe infections, the requirement of hospital and intensive care unit (ICU) admissions, and deaths associated with COVID-19, if any, through the end of the study will be assessed. In the safety analysis, adverse events were coded using the MedDRA Version 24.1 coding system.

The T-cell mediated immune response against the SARS-CoV-2 spike glycoprotein was assessed after the *in vitro* peptide stimulation of peripheral blood mononuclear cells (PBMC) followed by interferon gamma enzyme-linked immune absorbent spot (IFN-γ ELISpot) and intracellular cytokine staining (ICS) (details of procedures are described in the appendix (pp.3 Supplementary material).

### Outcomes

The primary endpoints were humoral immunogenicity measured by changes in levels of neutralizing antibodies (PBNA) against the ancestral Wuhan-Hu-1 strain after the PHH-1V or the BNT162b2 boost, and the safety and tolerability of PHH-1V as a boost. The secondary endpoints were to compare changes in levels of neutralizing antibodies against different variants of SARS-CoV-2, the T-cell responses towards the SARS-CoV-2 spike glycoprotein peptides and to compare the changes in immunogenicity measured by wild type SARS-CoV-2 neutralisation test (VNA) only in a subset of subjects, 20% approximately of the total subjects included in the study. The exploratory endpoint was to assess the number of subjects with SARS-CoV-2 infections ≥14 days after PHH-1V booster.

### Statistical analysis

The sample size was calculated in accordance with the Food and Drug Administration (FDA) Guidance for Industry on Clinical Data Needed to Support the Licensure of Seasonal Inactivated Influenza Vaccines. Noninferiority for a new influenza vaccine product could be claimed if the upper bound of the two-sided 95% confidence interval (CI) surrounding the ratio of GMT for the control to investigational product does not exceed 1·5. Superiority is concluded if the upper bound of the 95% confidence interval of the ratio of GMTs (BNT162b2: PHH-1V) is below 1. Given uncertainty in the immune response and variability, the Phase IIb part of this study was planned with a reduced non-inferiority margin of 1·4 to ensure sufficient sample size for safety and immunogenicity assessments.

Considering these assumptions, and with a 2:1 randomisation ratio, group sample sizes of 301 and 151, respectively, would achieve 90% power to detect non-inferiority using a one-sided 2·5% significance level, two-sample t-test using a SDlog=0·45 for both treatments. Assuming a 25% withdrawal rate, a total of 602 subjects (401 in PHH-1V group, 201 in the BNT162b2 active control group) were previewed to be randomised in this study. When reaching the 602 participants, the hospitals already had additional appointments for vaccination of participants, and in agreement with HIPRA and the CRO, it was decided to allow the hospitals to complete the vaccinations already scheduled during one additional week approximately. Otherwise, the hospitals would have to cancel the vaccinations already scheduled. A total of 782 subjects were enrolled into the Phase IIb part of the study, stratified 2:1 in the following age groups: 18 to 64 years and equal or over 65 years.

The following analyses of populations were included in this study: Intention-to-treat (ITT) population, including all subjects who were randomly assigned to treatment, regardless of the subject’s treatment status in the study; modified intention-to-treat (mITT) population, consisting of all subjects in the ITT who met the screening criteria and received one vaccine dose (subjects who tested positive for COVID-19 within 14 days of the receiving study drug were excluded); mITT3(98) population, including all subjects in the mITT without COVID-19 infections recorded via adverse event reporting prior to their Day 98 visit date; immunogenicity (IGP) population, including all subjects in the mITT who had a valid immunogenicity test result before receiving study drug and at least one valid result after dosing; per-protocol (PP), all subjects in the mITT who received one vaccine dose and had no major protocol deviations, and documented by the Sponsor prior to data base lock and unblinding that impact critical or key study data; safety (SP), comprised of all randomised subjects who received one vaccine dose, and were analysed according to the treatment received. The humoral immunogenicity analysis was done in both mITT and mITT3(98) populations, and the T-cell response from a subset of the total subjects included in three of the study sites. The safety analysis was conducted in the safety population and the assessment of COVID-19 cases in the PP population.

The humoral immunogenicity analyses tested the following hypotheses to show non-inferiority of PHH-1V when compared to the BNT162b2 booster vaccine: i) Null hypothesis, H_0_: the ratio of the GMTs (BNT162b2: PHH-1V) exceeds the non-inferiority margin (NI_m_); equivalently, the difference in log (GMT) exceeds log (NI_m_); ii) Alternative hypothesis, H_1_: the ratio of GMTs is below NI_m_; equivalently the difference in log (GMT) is less than log (NI_m_). The NI_m_ established for this study was 1·4, whereby the upper bound of the 95% confidence interval (95% CI) had to be lower than this value to accept the null hypothesis and was defined for each endpoint separately.

For the immunogenicity analysis, concerning the log_10_-transformed PBNA and VNA data on neutralising antibody titres against the SARS-CoV-2 variants and Wuhan-Hu-1 strain as well as the total binding antibodies data, linear mixed effects models were employed. In these models, the treatment group, the age group, the visit (baseline and day 14 for VNA and day 28 and 98 for PBNA) and the treatment-by-visit interaction were included as fixed effects, while the site and the subject-nested-to-site factors were included as random effects in random intercept models. In all models, a compound symmetry variance-covariance matrix structure was used, and restricted maximum likelihood was employed for parameter estimation. Denominator degrees-of-freedom were calculated using the Kenward-Roger’s approximation. Validity of the models was assessed graphically (i.e., quantile-quantile plots, residuals plots). For pairwise and interaction contrasts, estimated marginal means were calculated and compared.

Similarly, linear mixed effects models were employed for the analysis of the T-cell response data (ELISpot and ICS). Importantly, the proportions of positive cells were transformed using the angular (arcsine-square root) transformation. In these models, the visit, the treatment, the peptide pool stimulus, and the three-way interaction were included as fixed effects, while the subject identifier was included as a random effect. In all models, a heterogeneous first order autoregressive variance-covariance matrix structured was employed, and restricted maximum likelihood was used for parameter estimation. Validity of the models was assessed graphically (i.e., quantile-quantile plots, residuals plots). For pairwise and interaction contrasts, estimated marginal means were calculated and compared. To analyse the antibody neutralisation titre against SARS-CoV-2 (Wuhan-Hu-1, Beta, Delta, and Omicron BA.1) as measured by ID_50_ by PBNA or VNA, a mixed effects model for repeated measures (MMRM) was carried out on log transformed data. Summary statistics for the log_10_ transformations for each individual sample were calculated based on the log_10_-transformed titres at baseline, on day 14, day 28 and day 98, and are presented for the mITT population.

The GMFR analysis was performed for participants who, after a booster dose, had a ≥4-fold change in binding antibodies titre from baseline to day 14, day 28 and day 98 (responders), summarised by the number and proportion of responders along with an exact 95% Clopper-Pearson CI for the proportion.

For the cellular immunogenicity analysis, a MMRM of T-cell data (IFN-γ Spot-Forming Units [SFU]/10^6^ PBMCs) was employed, using the angular-transformed proportions as the response variable.

For the incidence of COVID-19 data are shown as the number of events and percentage of participants affected in the PP population. An exact 95% Clopper-Pearson CI for the proportion of each endpoint was also presented.

This trial is registered at ClinicalTrials.gov, NCT05142553.

## Results

From 15 November 2021, 862 participants were screened, of whom, 782 adults were randomly assigned to PHH-1V (n=522) or BNT162b2 (n=260) booster vaccine groups (Figure 1). A total of 504 participants in the PHH-1V group and 248 participants in the BNT162b2 group were included in the mITT population. For the SP population, 513 and 252 participants were included in the PHH-1V and BNT162b2 groups, respectively. Five hundred and three subjects were included in the PHH-1V group and 246 participants in the BNT162b2 group (IGP population). Since this trial is *ongoing*, the PP population has not yet been defined as protocol deviations are still being assessed. Baseline characteristics were well balanced between the two groups, where the median age of participants was 42 years (range 19–76 years; p=0·65), 484 (63·3%) were female (p=1·00) and 755 (98·7%) identified as White (p=1·00; Table 1).

**Figure 1:**
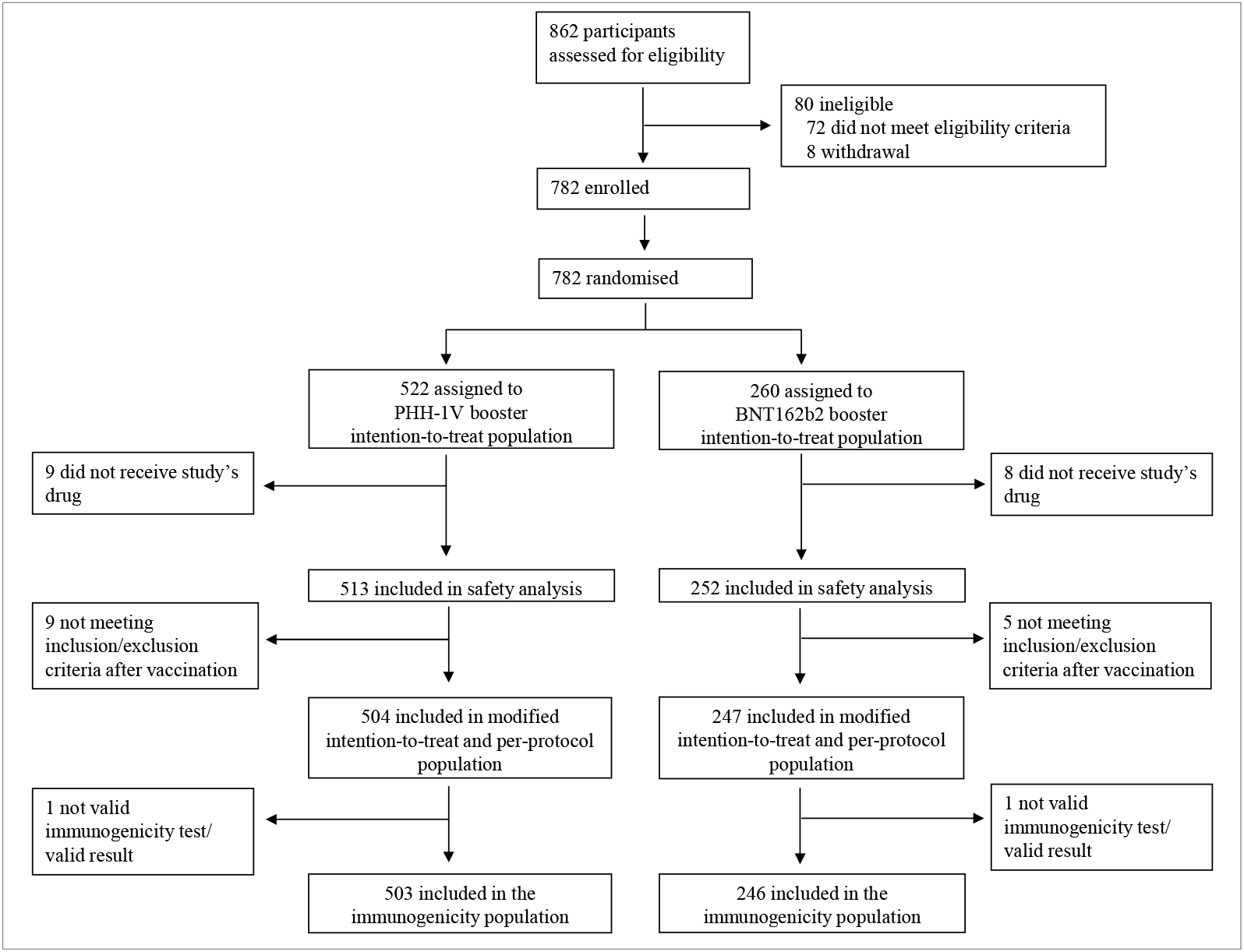
Trial Profile. *PHH-1V=PHH-1V vaccine, HIPRA. BNT162b2= BNT162b2 vaccine, Pfizer–BioNTech*.

**Table 1:** Baseline characteristics of participants included in the analysis.

|  | PHH-1V | BNT162b2 | Total | Confidence Interval | *p* value |
| --- | --- | --- | --- | --- | --- |
| **Number enrolled** | 513 (100·0) | 252 (100·0) | 765 (100·0) |  |  |
| **Age, years** |  |  |  |  |  |
| All age groups, median (range) | 42·0 (19·0-76·0) | 40·0 (20·0-74·0) | 42·0 (19·0-76·0) | 0·50 [-1·72,2·72] † | 0·65 |
| 18-65 | 475 (92·6) | 234 (92·9) | 706 (92·7) |  |  |
| ≥ 65 | 38 (7·4) | 18 (7·1) | 56 (7·3) | 0·96 [0·53,1·73] ‡ | 1·00 |
| **Sex** |  |  |  |  |  |
| Female | 325 (63·4) | 159 (63·1) | 484 (63·3) |  |  |
| Male | 188 (36·6) | 93 (36·9) | 281 (36·7) | 0·99 [0·72,1·37] ‡ | 1·00 |
| **Race** |  |  |  |  |  |
| White | 505 (98·4) | 250 (99·2) | 755 (98·7) |  |  |
| Asian | 4 (0·8) | 1 (0·4) | 5 (0·6) |  | 1·00 |
| American Indian or Alaska native | 2 (0·4) | 0 (0·0) | 2 (0·3) |  |  |
| Other | 2 (0·4) | 1 (0·4) | 3 (0·4) |  |  |
| **Ethnicity** |  |  |  |  |  |
| Hispanic or Latino | 170 (33·1) | 77 (30·6) | 247 (32·3) | 1·13 [0·8,1·56] ‡ | 0·51 |
| Not Hispanic or Latino | 343 (66·9) | 175 (69·4) | 518 (67·7) |  |  |
| **BMI, median (range)** | 24·03 (16·98-39·78) | 23·73 (17·90-39·86) | 23·92 (16·98-39·86) | 0·16 [-0·47,0·79] † | 0·61 |
| **Time between second dose and booster,** |  |  |  |  |  |
| *n* * | 446 | 218 | 664 | - | - |
| days, median (range) | 296·5 (162, 325) | 296·0 (182, 318) | 296·0 (162, 325) |  |  |

Data are *n* (%) unless otherwise specified. Confidence intervals are shown as † mean difference, 95% CI for the comparison of basal continuous variables or ‡ Odds Ratio, 95% CI for the comparison of dichotomous variables between groups. \*Participants who did not have the complete date in the second or third dose and those in which the subtraction “3rd dose -2nd dose” was negative were excluded from the analysis. BMI=body-mass index.

The GMT and GMFR of neutralizing antibodies determined by PBNA on day 14, 28 and 98 post-booster are shown in Table 2 and Figure 2A for the mITT population. For the primary endpoint of humoral immunogenicity measured by changes in levels of neutralizing antibodies (PBNA) against the ancestral Wuhan-Hu-1 strain, the GMT on day 14 was 1965·79 (95% CI: [1712·26, 2256·85]) for the PHH-1V group and 3309·29 (95% CI [2811·97, 3894·56]) for the BNT162b2 group, with a GMT ratio of 1·68 (95% CI [1·44, 1·97]; p<0·0001), which could not demonstrate non-inferiority of the PHH-1V vaccine to the BNT162b2 vaccine in immune response. The same was observed for day 28, with a GMT ratio of 1·31 (95% CI [1·12, 1·54]; p=0·0007). However, on day 98, the GMT ratio was 0·86 (95% CI [0·60, 1·22]; p=0·40), which shows non-inferiority of the PHH-1V vaccine to the BNT162b2 vaccine immune response. The GMFR data in Table 2 confirm the observed for GMT. A booster vaccination with PHH-1V could not demonstrate non-inferiority for Wuhan-Hu-1 on day 14 (ratio 1·74, 95% CI [1·44, 2·09]; p<0·0001) and day 28 (ratio 1·35, 95% CI [1·13, 1·63]; p=0·0013), but it did so for day 98 (ratio 0·89, 95% CI [0·61, 1·28]; p=0·52). These humoral immunogenicity results were additionally confirmed in the mITT3(98) population (excluding those subjects who reported COVID-19 infections trough day 98; Supplementary Table 1), where non-inferiority was demonstrated in day 98, with a GMT and a GMFR ratio of 0·84 [0·59, 1·20] (p=0·34) and 0·92 [0·63, 1·34] (p=0·65), respectively.

**Table 2:**
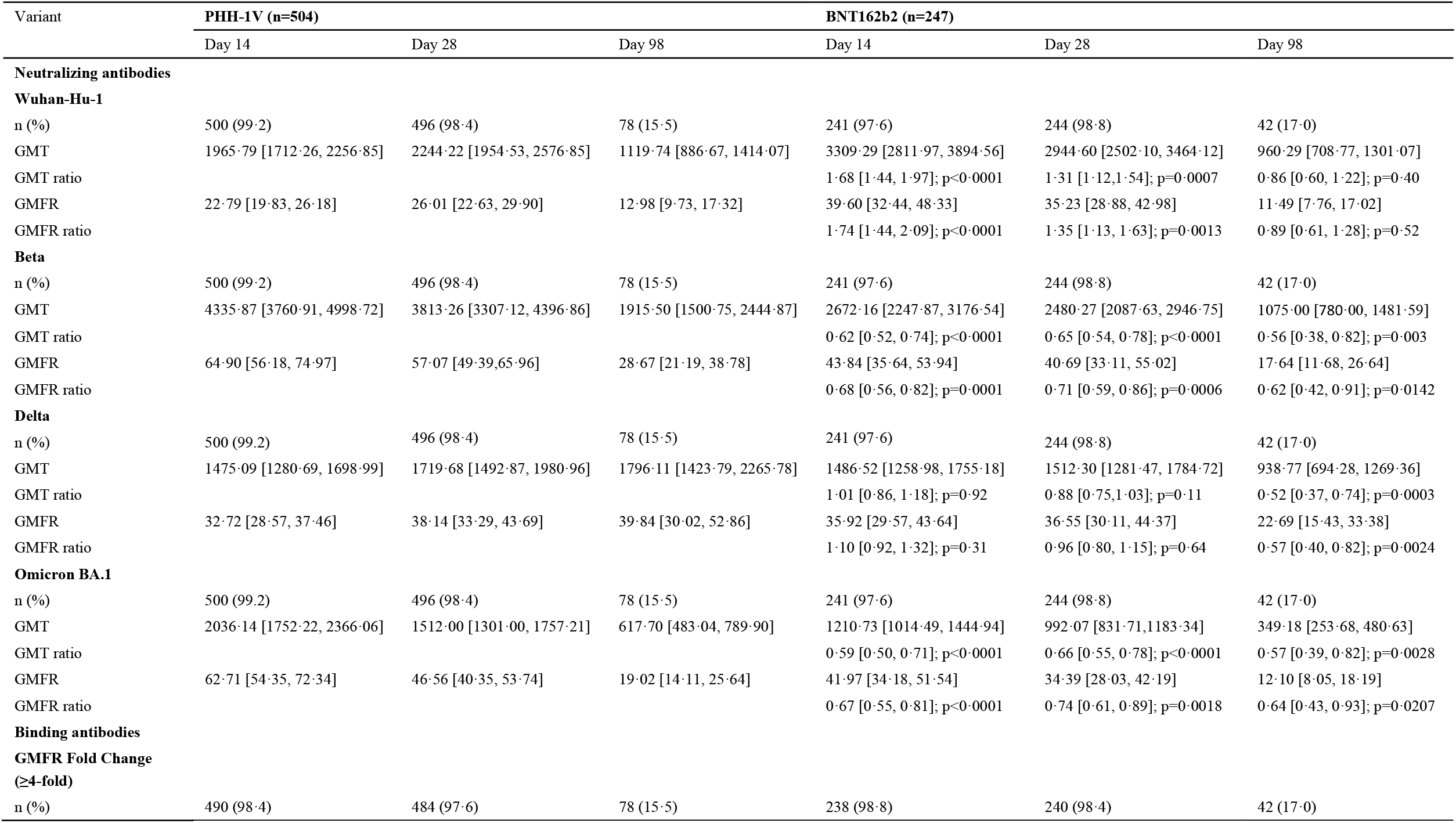
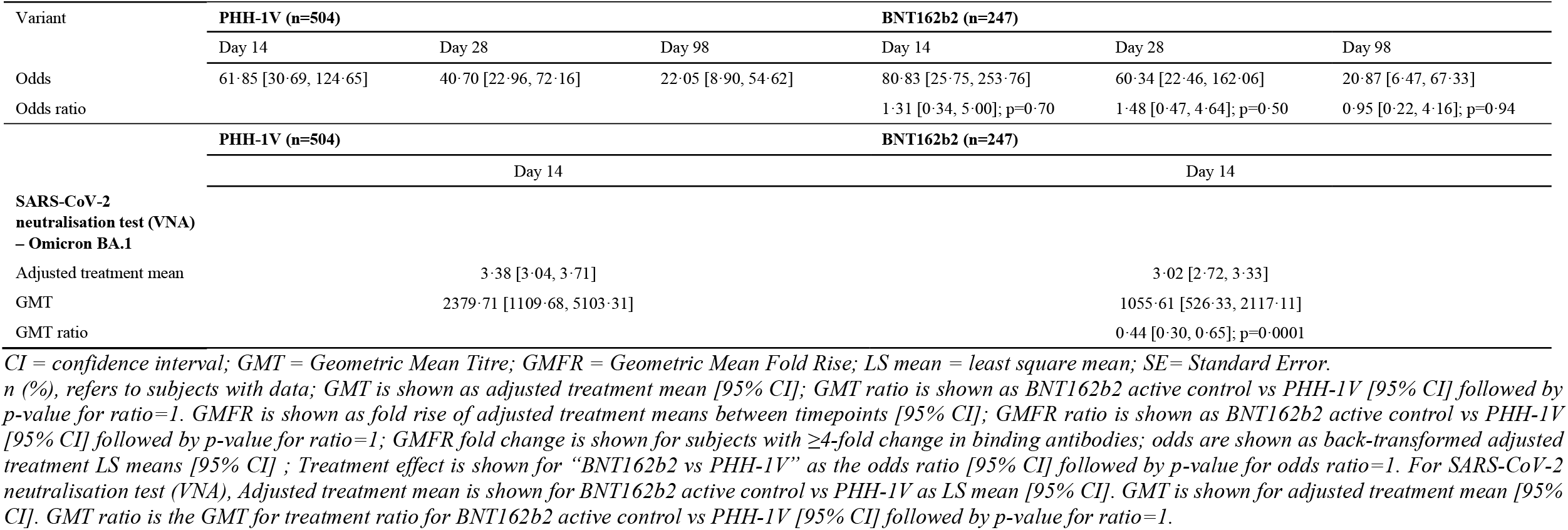
Analysis of neutralizing and binding antibodies against SARS-CoV-2 variants on days 14, 28 and 98 post-vaccination boost.

**Figure 2:**
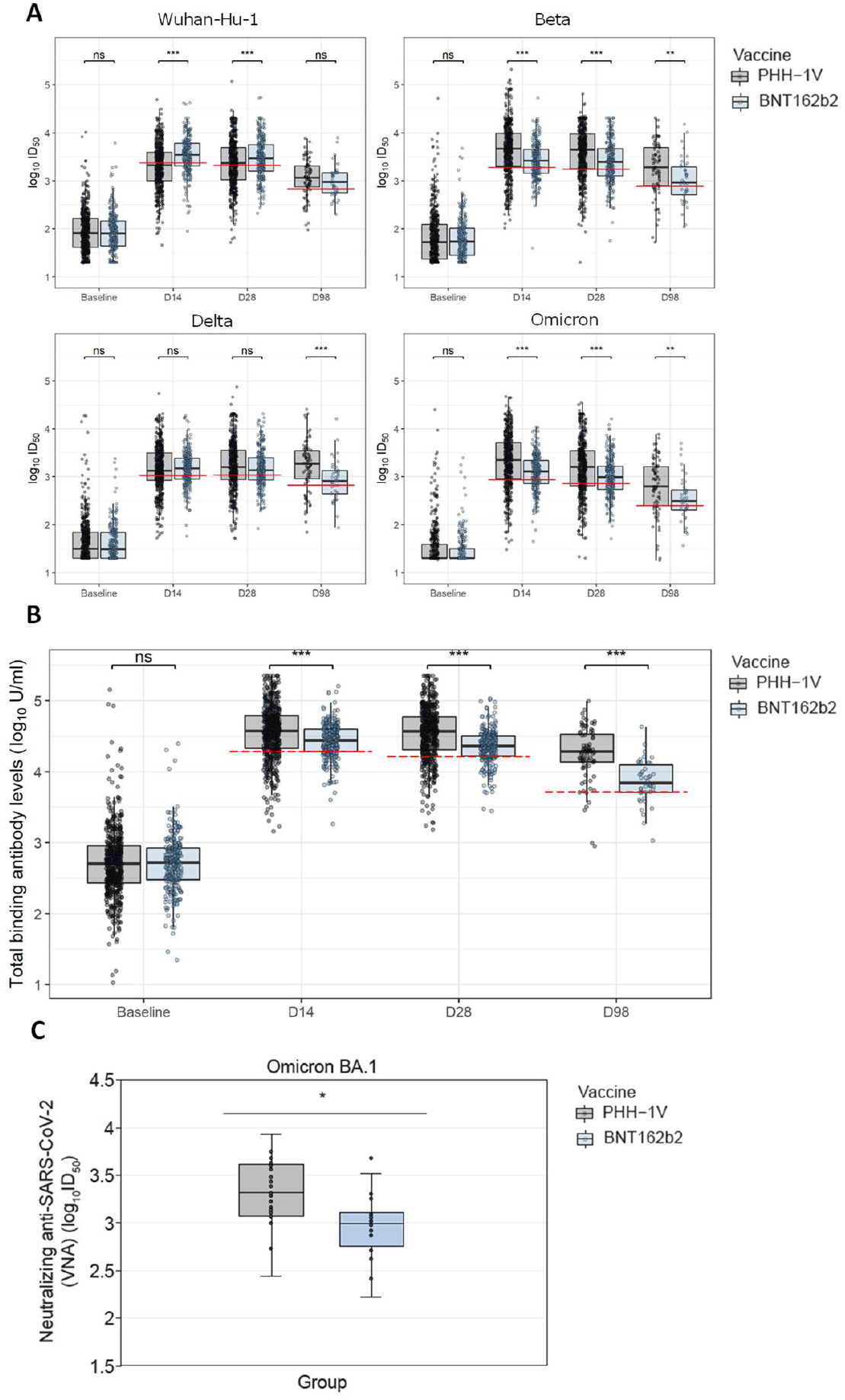
Analysis of the antibody response to PHH-1V vaccination. **(A)** Neutralizing antibody responses against multiple SARS-CoV-2 variants by PBNA in the mITT population. Comparison of neutralizing antibody titres at baseline, day 14, day 28 and day 98, between the PHH-1V and BNT162b2-vaccinated groups. **(B)** SARS-CoV-2 binding antibodies titre. Comparison of total binding antibody titres at baseline, day 14, day 28 and day 98, between the PHH-1V and BNT162b2-vaccinated groups. The analysis has been performed considering only responder subjects (≥4 fold-change in binding antibodies titre from baseline to day 14, day 28 or day 98). **(C)** SARS-CoV-2 neutralisation assay (VNA) for the Omicron variant (B.1.1.529). VNA assay was performed with serial dilutions of heat-inactivated serum samples from individuals receiving a PHH-1V or BNT162b2 boosting vaccine on day 14. VNA titres are plotted as ID_50_ (the reciprocal dilution inhibiting 50% of the cytopathic effect). *p-value for ratio=1, BNT162b2 active control *vs* PHH-1V; p=0·0001. Adjusted treatment mean is shown for VNA analysis for BNT162b2 active control (n=24) *vs* PHH-1V (n=34) as LS mean (95% CI). GMT is shown for adjusted treatment mean (95% CI). GMT ratio is the GMT for treatment ratio for BNT162b2 active control vs PHH-1V (95% CI) followed by p-value for ratio=1. Boxplots with grey dots and blue dots refer to PHH-1V and BNT162b2, respectively. The red dashed lines shown in the boxplots 2A and 2B indicate the non-inferiority limit, calculated as the GMT value of BNT162b2 divided by 1·4. Statistically significant differences are shown as * for *p* ≤ 0·05; ** for *p* ≤ 0·01; *** for *p* ≤ 0·001. Non-significant comparisons have been indicated with “ns”.

Concerning the primary endpoint of safety and tolerability, complete data for the vaccination diary are shown in the appendix up to day 7 post-boost (Supplementary Table 2). Briefly, the most frequent solicited local reactions on day 1 were pain (51·1% and 69·8% for the PHH-1V and the BNT162b2 group, respectively) and tenderness (48·5% for the PHH-1V and 63·5% for the BNT162b2 group) (Supplementary Table 2). The most frequent post-vaccination solicited systemic adverse events, for the PHH-1V and the BNT162b2 groups on day 1, respectively, were fatigue (16·0% and 35·3%), headache (14·2% and 27·8%), muscle pain (11·7% and 29·4%), and fever (0·6% and 7·1%) (Supplementary Table 2).

Overall, 458 (89·3%) subjects in the PHH-1V group and 238 (94·4%) subjects in the BNT162b2 group experienced at least one adverse event (p=0·0219) through day 28 after administration. Specifically, events of mild intensity were reported by 66·7% and 57·9% of subjects in the PHH-1V and BNT162b2 groups, respectively (p=0·02; Figure 3 and Supplementary Table 3). The most frequent adverse events were injection site pain (79.7% and 89·3%; p=0·0010), headache (31·2 and 40·1%; p=0·0190) and fatigue (27·5% and 42·1%; p=0·001) for the PHH-1V and the BNT162b2 groups, respectively (Figure 3 and Supplementary Table 3). Treatment-related adverse events were reported in 434 subjects (1384 events, 84·6%) in the PHH-1V group and 231 subjects (975 events, 91·7%) in the BNT162b2 group (p=0·0061). No deaths were reported in the study in either of the PHH-1V or BNT162b2 group and one SAE was reported for the PHH-1V group (0·2%, p=1·0; Supplementary Table 3).

**Figure 3:**
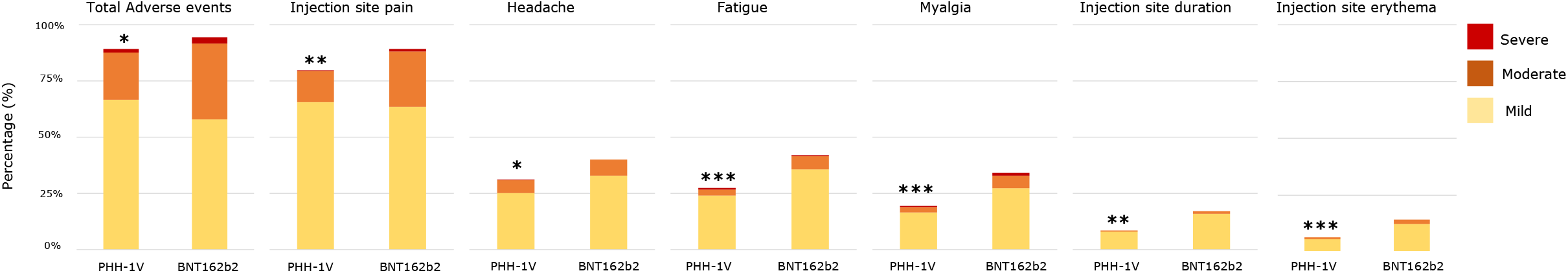
Adverse Events by MedDRA Preferred Term (PT) by treatment group through day 28 after booster administration reported in ≥10·0% of Overall Subjects. Data are shown as the percentage of subjects in relation to the safety population (PHH-1V, N=513 and BNT162b2, N=252), indicating the intensity of adverse events (mild, moderate and severe). If a subject experienced more than one adverse event, the subject is counted once for each PT choosing the most severe. PTs are ordered in decreasing frequency of the total number of subjects with each adverse event in PHH-1V group. \**p*<0·05; \*\**p*<0·001; \*\*\**p*<0·0001 (data from Supplementary Table 3).

For the secondary endpoint of humoral immunogenicity against different variants of SARS-CoV-2, the GMT for the beta variant on day 14 was 4335·87 (95% CI [3760·91, 4998·72]) in the PHH-1V group and 2672·16 (95% CI [2247·87, 3176·54]) in the BNT162b2 group, with a GMT ratio of 0·62 (95% CI [0·52, 0·74]; p<0·0001), indicating a superiority of the PHH-1V vaccine. The GMT ratios of 0·65 (95% CI [0·54, 0·78]; p<0·001) and 0·56 (95% CI [0·38, 0·82]; p=0·003) on days 28 and 98, respectively, confirm the superiority of the PHH-1V for the Beta variant. Regarding the Delta variant, the GMT on day 14 was 1475·09 (95% CI [1280·69, 1698·99]) and 1486·52 (95% CI [1258·98, 1755·18]) for the PHH-1V and BNT162b2 groups, respectively, with a GMT ratio of 1·01 (95% CI [0·86, 1·18]; p=0·92), demonstrating non-inferiority of the PHH-1V vaccine to BNT162b2. On days 28 and 98, the GMT ratio of 0·88 (95% CI [0·75, 1·03]; p=0·11) and 0·52 (95% CI [0·37, 0·74]; p=0·0003) indicates an inconclusive result and a superiority of the PHH-1V vaccine for the Delta variant, respectively. For the Omicron BA.1 variant, the PHH-1V and the BNT162b2 boosted groups presented a GMT of 2036·14 (95% CI [1752·22, 2366·06]) and 1210·73 (95% CI [1014·49, 1444·94]), respectively, and a GMT ratio of 0·59 (95% CI [0·50, 0·71]; p<0·0001) on day 14, thus demonstrating superiority of the PHH-1V vaccine to the BNT162b2 vaccine against the Omicron BA.1 SARS-CoV-2 variant. This result is further confirmed on days 28 and 98, with a GMT ratio of 0·66 (95% CI [0·55, 0·78]; p<0·001) and 0·57 (95% CI [0·39, 0·82]; p=0·0028), respectively. All results for GMT and GMT ratios were confirmed in the mITT3(98) population (Supplementary Table 1), except for the inconclusive result for the Delta variant in day 28, which demonstrated a non-inferiority when compared to BNT162b2.

The GMFR ratios concerning the different assessed variants of SARS-CoV-2, the data in Table 2 confirm the results observed for GMT on day 14. A booster vaccination with PHH-1V provided non-inferiority for the Delta variant (ratio 1·10, 95% CI [0·92, 1·32]; p=0·31), and elicited statistically significant higher levels of neutralizing antibodies for the Beta (ratio 0·68, 95% CI [0·56, 0·82]; p=0·0001) and Omicron BA.1 (ratio 0·67, 95% CI [0·55-0·81]; p<0·0001) variants, thus, demonstrating superiority of the PHH-1V booster against Beta and Omicron BA.1 variants. On day 28, a booster vaccination with PHH-1V demonstrates non-inferiority for Delta variant (ratio 0·96, 95% CI [0·80, 1·15]; p=0·64), and a superiority of PHH-1V was observed for Beta and Omicron BA.1 variants (ratios of 0·71, 95% CI [0·59, 0·86]; p=0·0006 and 0·74, 95% CI [0·61, 0·89]; p=0·0018, respectively). On day 98 post-boost, the GMFR ratio for the Beta (ratio 0·62, 95% CI [0·42, 0·91]; p=0·0142), Delta (ratio 0·57, 95% CI [0·40, 0·82]; p=0·0024), and Omicron BA.1 (ratio 0·64, 95% CI [0·43, 0·93]; p=0·0207) variants demonstrates superiority of the PHH-1V vaccine to the BNT162b2 vaccine against these variants. These results were further confirmed when the data analysis was performed with the mITT3(98) population (Supplementary Table 1).

The percentage of participants with a ≥4-fold change in binding antibodies in the PHH-1V and BNT162b2 vaccine groups were similar, 98·4% (n=490) and 98·8% (n=238) respectively, on day 14 post-boost (Table 2). Accordingly, there were non-significant differences between the odds of the two groups, with an odds ratio of 1·31 (95% CI [0·34, 5·00]; p=0·70). The data concerning day 28 and 98 after booster administration reinforce this result, with non-significant differences in the odds between treatment groups (D28 odds ratio 1·48, 95% CI [0·47, 4·64]; p= 0·50 and D98 odds ratio 0·95, 95% CI [0·22, 4·16]; p= 0·94). Nevertheless, the quantitative comparison of the total binding antibody titres between treatments reflects that vaccination with PHH-1V induced higher antibody titres on days 14, 28 and 98 compared with the BNT162b2 booster dose (p<0·0001; Figure 2B). These results indicate that a similar percentage of subjects respond to both vaccines with a ≥4-fold change in binding antibodies, although the subjects vaccinated with PHH-1V reach higher titres compared with those immunized with BNT162b2. Interestingly, quantitative results on day 98 show that the decrease in the titre in the PHH-1V arm is not as pronounced as in the BNT162b2 arm, with geometric mean fold reduction ratio from day 14 to day 98 of 1·87 (95% CI [1·35, 2·60]; p<0·0001), suggesting a greater maintenance of the long-term antibody titre (Figure 2B).

The results for the SARS-CoV-2 neutralisation test (VNA) for the Omicron BA.1 variant on day 14 show a GMT for treatment ratio for BNT162b2 active control *vs* PHH-1V of 0·44 (95% CI [0·30, 0·65]) followed by *p* value for ratio=1 (p=0·0001; Figure 2C and Table 2). The baseline characteristics of this subset of 58 participants is shown in Supplementary Table 4.

For the secondary endpoint of assessing T-cell responses towards the SARS-CoV-2 spike glycoprotein peptides, participants in the PHH-1V group showed a significant increase of IFN-γ producing lymphocytes upon *in vitro* re-stimulation with SARS-CoV-2 spike peptide pools SA and RBD (4 pools covering Wuhan-Hu-1, Alpha, Beta, and Delta variants; appendix pp. 3 Supplementary material). A significant activation of CD4^+^ T-cells expressing IFN-γ was observed for the PHH-1V group at 2 weeks compared with baseline upon the stimulation with the RBD (Wuhan-Hu-1, Alpha, and Beta variants) and Spike SA peptide pool (Figure 4A; p<0·0001). For these variants, the increase rate in the percentage of CD4^+^ IFN-γ^+^ T-cells from baseline to 2 weeks was higher in the PHH-1V group compared to the BNT162b2 group (Wuhan-Hu-1, p=0·0097; Alpha, p=0·0030; Beta, p=0·0069). Regarding the CD8^+^ T-cells, the boost with the PHH-1V vaccine significantly increased the percentage of IFN-γ-expressing cells upon re-stimulation with RBD from SARS-CoV-2 Beta variant at week 2 compared to baseline (Figure 4B; p=0·0372). The BNT162b2 vaccine boost increased the percentage of CD8^+^ IFN-γ^+^ T-cells after re-stimulation with a spike peptide pool SA (p=0·0089) and SB (p=0·0018) at week 2 compared with baseline, showing higher values in the increase rate than the PHH-1V boost (p=0·0147 and p=0·0248, respectively). Moreover, the BNT162b2 group increased the percentage of CD8^+^ IL-4^+^ T-cells after re-stimulation with the SB spike peptide pool at week 2 compared with baseline (p=0·0123).

**Figure 4:**
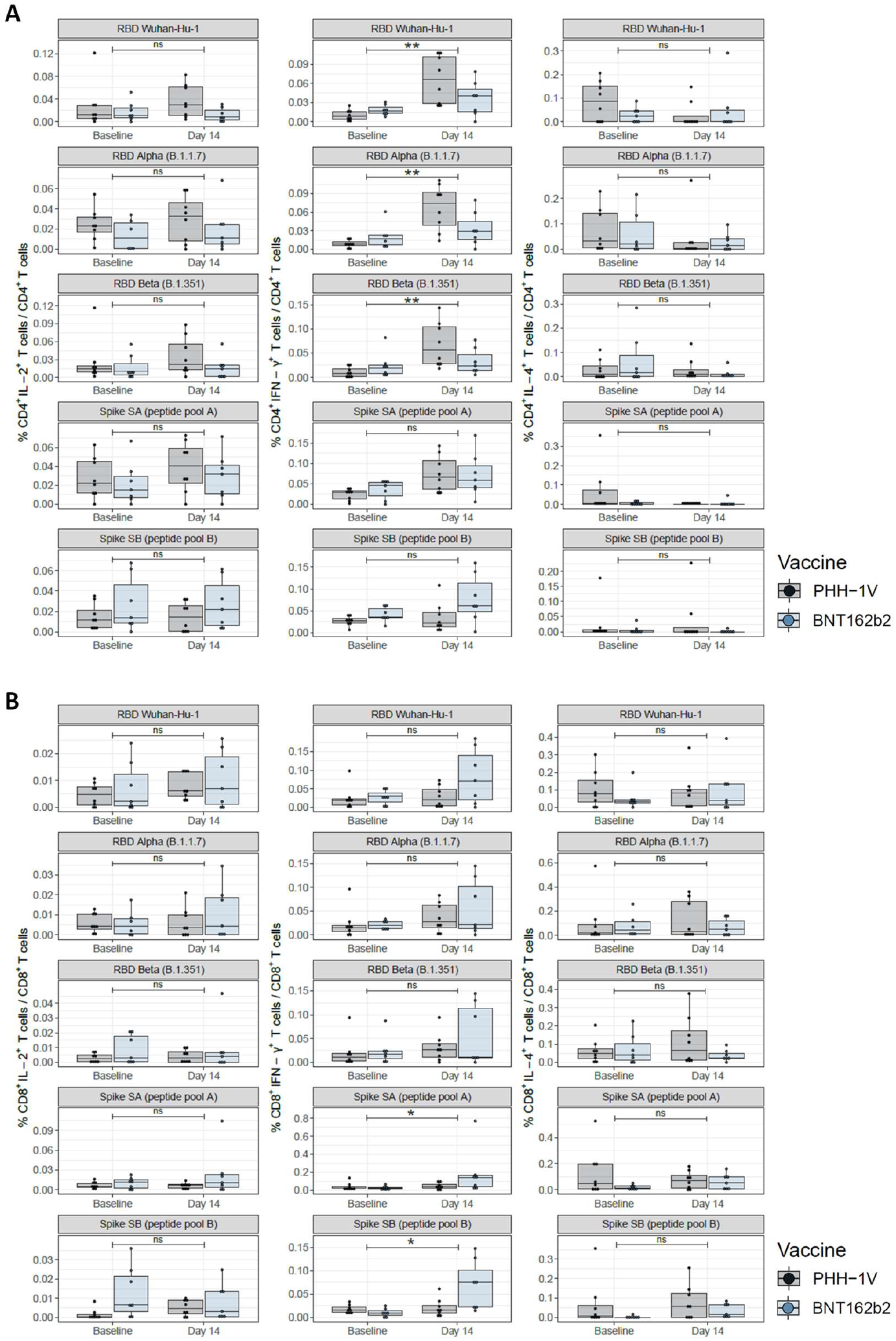
Characterization of T-cell responses in PBMCs from groups receiving a heterologous boosting vaccination with PHH-1V (in grey) or BNT162b2 (in blue). CD4^+^ T (**A**) and CD8^+^ T (**B**) cells were characterized by intracellular cytokine staining (ICS) of interleukin-2 (IL-2^+^; left panel), interferon gamma (IFN-γ^+^; middle panel) and interleukin-4 (IL-4^+^, right panel). Boxes depict the median (solid line) and the interquartile range (IQR), and whiskers expand each box edge 1·5 times the IQR. Interaction contrasts have been displayed in the plots, comparing the increase rates over time between the two vaccination groups. Statistically significant differences are shown as * for *p* ≤ 0·05; ** for *p* ≤ 0·01; *** for *p* ≤ 0·001. Non-significant comparisons have been indicated with “ns”. *P-*values lower than 0·05 indicate that one of the vaccinated groups has experienced a stronger boost compared to the other. *RDB; receptor binding domain for the SARS-CoV-2 spike protein (ancestor Wuhan-Hu-1 strain); RDB B*.*1*.*1*.*7 (Alpha variant); RDB B*.*1*.*351 (Beta variant); Spike SA corresponds to 194 spike protein peptide pools overlapping the S1-2016 to S1-2196 region of the Spike protein; Spike SB corresponds to 168 spike protein peptide pools overlapping the S1-2197 to S2-2377 region of the Spike protein*.

Concerning the exploratory endpoint of SARS-CoV-2 infections ≥14 days post-booster administration, at the data cut-off date of the report the percentage of patients who reported experiencing COVID-19 (n=82) remained similar in the PHH-1V (10·14%) and BNT162b2 (11·90%) groups (OR 0·83, 95% CI [0·51, 1·36]; p= 0·45). None of the subjects developed severe COVID-19 and, consequently, no hospital or intensive care unit (ICU) admission were required.

## Discussion

Our interim results from the Phase IIb HH-2 trial show that PHH-1V as a heterologous boost elicits a non-inferior neutralizing antibody response to SARS-CoV-2 Delta variant, a superior neutralizing antibody response against Beta variant and, most importantly, a superior neutralizing antibody response against the Omicron variant of concern (sublineage BA.1, predominant during the development of the trial) at 14 days post-vaccination when compared to the BNT162b2 boost. Although the PHH-1V as a booster could not demonstrate non-inferiority to BNT162b2 against the initial Wuhan-Hu-1 SARS-CoV-2 strain on days 14 and 28, it provided non-inferiority on day 98. Moreover, on day 98, PHH-1V vaccine showed superiority for the Delta variant. These results suggest a lower decrease in the neutralizing antibody titres after the PHH-1V boost when compared to BNT162b2. Accordingly, given the very high levels of community transmission observed for the currently circulating Delta and Omicron variants, neutralisation antibody response against these variants is of paramount importance. No significant differences were observed between the groups for those participants with ≥4-fold change in binding antibodies up to 98 days after boost administration, although the PHH-1V induced higher titres compared with the BNT162b2 boost. Our results have also shown that PHH-1V as a booster dose induces a significant increase of CD4^+^ and CD8^+^ T-cells expressing IFN-γ on day 14. Concerning the safety profile, PHH-1V group had less percentage of adverse events compared with BNT162b2 group, with most of mild intensity, and similar non-severe COVID-19 cases.

For the trial’s primary endpoint of humoral immunogenicity against the ancestral Wuhan-Hu-1 strain, the BNT162b2 booster elicited a superior neutralizing antibody response on days 14 and 28, with a non-inferiority demonstrated for PHH-1V only at Day 98. This result may be explained by the PHH-1V vaccine being based on a fusion heterodimer consisting of the spike RBD sequence from the SARS-CoV-2 Beta and Alpha variants, while the BNT162b2 vaccine encodes the SARS-CoV-2 full-length spike protein of Wuhan-Hu-1^30^.

For the primary endpoint of safety and tolerability, the percentage of subjects with local and systemic reactions, total adverse effects and treatment-related adverse events were lower in the PHH-1V group compared to the BNT162b2 group. No deaths were reported in the study in either of the PHH-1V or BNT162b2 group and one SAE was reported, which was resolved and found to be nonrelated to the vaccination. As of day 98, a total of 82 SARS-CoV-2 cases were reported, and no statistically significant differences were observed between the PHH-1V booster and the active control, BNT162b2. In addition, no severe cases of COVID-19 were reported in any of the vaccinated groups. Of note, this study took place temporarily during a period of high incidence of COVID-19, namely the currently circulating Delta and Omicron (BA.1 variant) waves in Spain. Moreover, most of participants in the study were healthcare workers, thus with a higher infection exposition compared to the general population.^31^

When assessing the secondary endpoint of humoral immunogenicity against different variants of SARS-CoV-2, remarkably, PHH-1V was able to sustain a good neutralising ability against the Omicron BA.1 SARS-CoV-2 variant despite the heavily mutated spike protein of this variant, many of them located in the RBD.^32^ This fact strongly supports the high humoral immunogenicity of the PHH-1V RBD-based candidate against a wide range of potential new mutations since the PHH-1V antigen comprises key mutations that are also present in the Omicron BA.1 variant, as well as in many other SARS-CoV-2 variants. In addition, our results in humoral immunogenicity were confirmed when all the subjects who reported COVID-19 infections were excluded of the data analysis, thus eliminating the possible bias of an uneven distribution of infection-induced immunogenicity.

For the secondary endpoint of T-cell-mediated response determined by ELISpot, the results indicate that the PHH-1V boost, after a primary vaccination with BNT162b2, increases the cellular immune response after the *in vitro* re-stimulation. The ICS data for T-cell response characterization demonstrates that the booster immunization with the PHH-1V vaccine induces the activation of CD4^+^ T-cells expressing IFN-γ upon re-stimulation with pools of RBD peptides from different variants. Interestingly, this response is stronger compared to those subjects boosted with BNT162b2 vaccine, indicating that the PHH-1V heterologous boost is more efficient than the homologous boost in the activation of the CD4^+^ T-cell memory previously induced by the primary vaccination protocol. As no IL-4 expression was detected in the activated CD4^+^ T-cells after the *in vitro* re-stimulation, the ICS results suggest that the PHH-1V booster induces a Th1-biased T-cell response. The detection of the IFN-γ expression by the ELISpot assay confirms the Th1-biassed T-cell response upon boost immunization with the PHH-1V vaccine. Avoiding a Th2-biased immune response is important as it has been related with ineffective vaccines that induce vaccine-associated enhanced respiratory disease (VAERD) after subsequent infection.^33^ Moreover, the heterologous boost with the PHH-1V vaccine has proven to induce the activation of CD8^+^ T-cells expressing IFN-γ. The T-cell response is crucial to confer protection from COVID-19 severe disease^4^, and these results show the potential ability of PHH-1V not only to fight against the virus with neutralizing antibodies but also with cellular immunity specifically to destroy infected cells.

When comparing PHH-1V with other vaccines, the CoVLP is a plant-produced virus-like particle (VLP) vaccine under development, and the CoV2 preS dTM is an authorised recombinant protein antigen vaccine based on an S-protein sequence from the Wuhan-Hu-1 (D614) reference strain.^34,35^ Both candidate vaccines have been initially tested without adjuvant, but elicited better immunogenicity results with the AS03 adjuvant (GlaxoSmithKline Vaccines, Rixensart, Belgium), and are now called CoVLP-AS03 and CoV2 preS dTM-AS03, respectively. In addition, their phase II results indicate that they are well tolerated and have favourable neutralizing antibody titres, with both being stable at refrigerator temperatures. Unlike PHH-1V, which is being tested as a booster only, these vaccines were tested for primary (2-dose) vaccination schedules, thereby, without non-inferiority assessments with already approved vaccines. The NVX-CoV2373 subunit vaccine was also tested for a 2-dose vaccination schedule, eliciting lower IgG levels compared to the BNT162b2 and mRNA-1273 mRNA vaccines and a poor induction of spike-specific CD8^+^ T-cells.^36,37^ These data contrast with those reported in the present study for PHH-1V. However, the NVX-CoV2373 induced spike-specific CD4^+^ T-cells to all tested VOCs (Wuhan-Hu-1, Alpha, Beta, Delta and Omicron BA.1 and BA.2).^37^ The CoVLP-AS03, CoV2 preS dTM-AS03, and NVX-CoV2373 vaccines are now also being considered as booster doses for primary vaccination schedules.

The main limitations of this study are related to the changing epidemiological and social situation during the pandemics. Although it was initially proposed that neutralising antibodies by means of PBNA would have to be determined for Wuhan-Hu-1 strain and Alpha, Beta and Delta SARS-CoV-2 variants, the situation of the pandemics during the study made us change the proposal and so Alpha was substituted for Omicron which was the relevant circulating variant at that time, adding a very important value to the study that has demonstrated the effective potential of the PHH-1V vaccine as a heterologous booster. However, pseudovirions against the more recent circulating SARS-CoV-2 variants, such as Omicron BQ.1.1 or XBB, for example, are not yet available to perform most of the assays presented in this article.

In conclusion, the HH-2 study demonstrates that the PHH-1V vaccine is safe and well tolerated, eliciting a strong neutralizing antibody response against all tested SARS-CoV-2 variants and Wuhan-Hu-1 strain. Although PHH-1V did not reach non-inferiority at day 14 and day 28 in neutralizing antibody response against the Wuhan-Hu-1 strain, it did so at day 98. The study specifically shows superiority against the Beta and Omicron BA.1 SARS-CoV-2 variants when compared to BNT162b2 homologous vaccine boost and non-inferiority against Delta on day 14. Importantly, the PHH-1V vaccine also allows for better sustained levels of antibodies over time (day 98). This demonstrates that PHH-1V can elicit protection against current circulating variants of concern and, most importantly, can anticipate protection against potentially new emerging variants. The PHH-1V vaccine also induces a strong and balanced T-cell response against SARS-CoV-2. At present, a phase III trial is ongoing where the inclusion criteria have been opened further and studies on the vaccine are continued. The interim results presented here provide data on the balance observed between safety and immunogenicity elicited by the PHH-1V boost. These data support PHH-1V vaccine as a valuable tool to the current authorized COVID-19 vaccines and as a booster dose in the vaccination campaigns.

## Supporting information

Corominas et al 2022_supplementary

## Data Availability

All data relevant to the study are included in the article or uploaded as supplementary information. Further data are available from the authors upon reasonable request and with permission of HIPRA S.A.

## Contributors

Veristat was responsible for managing the data. Authors contributed to the acquisition, analysis, and/or interpretation of data. All authors had full access to all the data, revised the manuscript critically for important intellectual content, approved the version to be published, and accepted responsibility for publication.

## Acknowledgements

Medical writing support was provided by Vanessa Chigancas at Dynamic Science S.L.U. (Evidenze Clinical Research, Madrid, Spain) during the preparation of this paper, and funded by HIPRA SCIENTIFIC, S.L.U.

We especially acknowledge the following members of Veristat, who contributed to the success of this trial. The following were responsible for study management, biostatistics, medical monitoring, data management, and database programming of the study: Robin Bliss, PhD, Emma Albacar, MPH, Nancy Hsieh, MPH, Montse Barcelo, MD, Mariska van der Heijden, MSc, Amy Booth, Edmund Chiu, Avani Patel, and Cesar Wong, MD.

We are grateful to Daniel Perez-Zsolt and Jordana Muñoz-Basagoiti for their outstanding contribution to VNA.

The authors would like to thank Silvia Marfil and Raquel Ortiz for technical assistance.

The authors thank Ruth Peña for technical assistance with sample management and ELISpot and Gabriel Felipe Rodriguez-Lozano for ELISpot database generation.

We would like to express our gratitude to Marina Machado, Ana Álvarez-Uría, Sara Rodríguez, M^a^ Jesús Pérez Granda, Juan Carlos López Bernaldo de Quirós, M^a^ Teresa Aldamiz, Francisco Tejerina, Cristina Díez, Iván Adán, Ana Mur, Félix García and Víctor Fernández for their tireless effort and contribution to this important public health clinical trial.

We would like to thank Glòria Pujol and Eduard Fossas for their assistance in the revision of the manuscript; Fiorella Gallo, Núria Fuentes and Miriam Oria for the ELISA analysis; Clara Panosa, Thais Pentinat and Ester Puigvert for their assistance in the production of the vaccine and Jordi Palmada and Eva Pol for carrying out manufacturing controls. And of course, we would like to especially thank all the HIPRA workers who in one way or another have contributed to making this project a reality.

The authors thank the members of the DSMB for their expertise and recommendations.

We are indebted to the HCB-IDIBAPS Biobank, integrated in the Spanish National Biobanks Network, for the biological human samples and data procurement.

Finally, we want to express our gratitude to all the volunteers for their time and effort. With their contribution they have enabled the generation of medical and scientific knowledge that will enable us to draw closer to the end of this pandemic.

## Research in context

### Evidence before this study

Despite the efforts of mass vaccination programs against SARS-CoV-2, the situation is far from being under control. Only 68·5% of the world population, and, more starkly, 24·6% in low-income countries have received at least one dose of a COVID-19 vaccine. Moreover, because immunity wanes over time after both vaccination and/or infection, booster immunisation is crucial to control the SARS-CoV-2 infection rate and avoid saturation of health services. Accordingly, access to second-generation vaccines with a broader variants scope and longer-lasting protection is required. To date, NVX-CoV2373 (Novavax) is the only recombinant spike protein-based vaccine approved by WHO and EMA, and more vaccines platforms are needed. One would consider recombinant protein-based to be advantageous SARS-CoV-2 vaccines presentations; they are stable at 2-8 °C, with good safety and stability profiles, and suitable for people with compromised immune systems. We also searched PubMed on July 11^th^, 2022, with no date or language restrictions, and using the search terms “SARS-CoV-2” and “protein-vaccine” in the Title/Abstract. Our search revealed the vaccines RelCoVax^®^, S-26019-b, CoV2 preS dTM-AS03, zf2001, CoVaccine HT, NARUVAX-C19, Advax-SM, the nanoparticle Q@NP-adjuvanted S-protein vaccine, and the TLR7/8 agonist-adjuvanted recombinant spike protein vaccine with published data in animal models, while the NVX-CoV2373, Spikogen^®^, S-26019-b, V-01, Nanocovax, RDB Vaccine (Abdala), CoV2 preS dTM-AS03 and MVC-COV1901 had publications for their registered clinical trials. In addition, we searched public databases, as the “global database of COVID-19 vaccinations” (https://ourworldindata.org/covid-vaccinations) to have an update in COVID-19 vaccines doses administered, and the “COVID-19 Vaccine Development and Approvals Tracker, 2020” (available at covid19.trackvaccines.org) to check approved and in development vaccines sorted by type. Consequently, we believe that PHH-1V, a SARS-CoV-2 vaccine based on a fusion heterodimer consisting of the spike RBD sequence from the SARS-CoV-2 Beta (B.1.351) and Apha (B.1.1.7) variants, comes as a safe and efficient option for heterologous boost administrations regimens considering the interim data reported in this article for the HH-2 trial.

### Added value of this study

PHH-1V as a heterologous boost elicits a non-inferior neutralizing antibody response to SARS-CoV-2 previous and currently circulating variants when compared to the BNT162b2 boost. Noteworthy, PHH-1V conferred a superior neutralizing antibody response against the Omicron variant of concern (sublineage BA.1) at 14-, 28- and 98-days post-vaccination. PHH-1V as a booster dose induced a significant increase of CD4^+^ and CD8^+^ T-cells expressing IFN-γ, considered a biomarker of successful vaccination resulting in the development of effective humoral response. The vaccination with PHH-1V did not induce a Th2-biased immune response, an important aspect previously associated with ineffective vaccines that induce vaccine-associated enhanced respiratory disease. The PHH-1V group had less percentage of adverse events compared with BNT162b2 group, most of mild intensity.

### Implications of all the available evidence

PHH-1V can elicit protection against current circulating SARS-CoV-2 variants of concern. This protein-based vaccine represents a safe, well tolerated, and efficient option for heterologous booster, improving the range of vaccines available to cover diverse global demands and reach those with none or low vaccine coverage.

## References

1. Salamanna F, Maglio M, Landini MP, Fini M. Body Localization of ACE-2: On the Trail of the Keyhole of SARS-CoV-2. Front Med (Lausanne) 2020; 7: 594495.

2. Yang Y, Du L. SARS-CoV-2 spike protein: a key target for eliciting persistent neutralizing antibodies. Signal Transduct Target Ther 2021; 6(1): 95.

3. Khoury DS, Cromer D, Reynaldi A, et al. Neutralizing antibody levels are highly predictive of immune protection from symptomatic SARS-CoV-2 infection. Nat Med 2021; 27(7): 1205–11.

4. GeurtsvanKessel CH, Geers D, Schmitz KS, et al. Divergent SARS-CoV-2 Omicron-reactive T and B cell responses in COVID-19 vaccine recipients. Sci Immunol 2022; 7(69): eabo2202.

5. Ramasamy MN, Minassian AM, Ewer KJ, et al. Safety and immunogenicity of ChAdOx1 nCoV-19 vaccine administered in a prime-boost regimen in young and old adults (COV002): a single-blind, randomised, controlled, phase 2/3 trial. Lancet 2021; 396(10267): 1979–93.

6. Stuart ASV, Shaw RH, Liu X, et al. Immunogenicity, safety, and reactogenicity of heterologous COVID-19 primary vaccination incorporating mRNA, viral-vector, and protein-adjuvant vaccines in the UK (Com-COV2): a single-blind, randomised, phase 2, non-inferiority trial. Lancet 2022; 399(10319): 36–49.

7. Zhu FC, Li YH, Guan XH, et al. Safety, tolerability, and immunogenicity of a recombinant adenovirus type-5 vectored COVID-19 vaccine: a dose-escalation, open-label, non-randomised, first-in-human trial. Lancet 2020; 395(10240): 1845–54.

8. Heinz FX, Stiasny K. Distinguishing features of current COVID-19 vaccines: knowns and unknowns of antigen presentation and modes of action. NPJ Vaccines 2021; 6(1): 104.

9. Fahrni ML, Ismail IA, Refi DM, et al. Management of COVID-19 vaccines cold chain logistics: a scoping review. J Pharm Policy Pract 2022; 15(1): 16.

10. Dolgin E. How protein-based COVID vaccines could change the pandemic. Nature 2021; 599(7885): 359–60.

11. McDonald I, Murray SM, Reynolds CJ, Altmann DM, Boyton RJ. Comparative systematic review and meta-analysis of reactogenicity, immunogenicity and efficacy of vaccines against SARS-CoV-2. NPJ Vaccines 2021; 6(1): 74.

12. Mathieu E, Ritchie H, Ortiz-Ospina E, et al. A global database of COVID-19 vaccinations. Nat Hum Behav 2021; 5(7): 947–53.

13. Puhach O, Adea K, Hulo N, et al. Infectious viral load in unvaccinated and vaccinated individuals infected with ancestral, Delta or Omicron SARS-CoV-2. Nat Med 2022.

14. Agrati C, Capone S, Castilletti C, et al. Strong immunogenicity of heterologous prime-boost immunizations with the experimental vaccine GRAd-COV2 and BNT162b2 or ChAdOx1-nCOV19. NPJ Vaccines 2021; 6(1): 131.

15. Andersson NW, Thiesson EM, Laursen MV, Mogensen SH, Kjær J, Hviid A. Safety of heterologous primary and booster schedules with ChAdOx1-S and BNT162b2 or mRNA-1273 vaccines: nationwide cohort study. Bmj 2022; 378: e070483.

16. Atmar RL, Lyke KE, Deming ME, et al. Homologous and Heterologous Covid-19 Booster Vaccinations. N Engl J Med 2022; 386(11): 1046–57.

17. Interim recommendations for heterologous COVID-19 vaccine schedules: interim guidance, 16 December 2021. CC BY-NC-SA 3.0 IGO. Geneva: World Health Organization, 2021.

18. Costa Clemens SA, Weckx L, Clemens R, et al. Heterologous versus homologous COVID-19 booster vaccination in previous recipients of two doses of CoronaVac COVID-19 vaccine in Brazil (RHH-001): a phase 4, non-inferiority, single blind, randomised study. Lancet 2022; 399(10324): 521–9.

19. Garg I, Sheikh AB, Pal S, Shekhar R. Mix-and-Match COVID-19 Vaccinations (Heterologous Boost): A Review. Infect Dis Rep 2022; 14(4): 537–46.

20. Jara A, Undurraga EA, Zubizarreta JR, et al. Effectiveness of homologous and heterologous booster doses for an inactivated SARS-CoV-2 vaccine: a large-scale prospective cohort study. Lancet Glob Health 2022; 10(6): e798–e806.

21. Kleanthous H, Silverman JM, Makar KW, Yoon IK, Jackson N, Vaughn DW. Scientific rationale for developing potent RBD-based vaccines targeting COVID-19. NPJ Vaccines 2021; 6(1): 128.

22. Kyriakidis NC, López-Cortés A, González EV, Grimaldos AB, Prado EO. SARS-CoV-2 vaccines strategies: a comprehensive review of phase 3 candidates. NPJ Vaccines 2021; 6(1): 28.

23. Barreiro A, Prenafeta A, Bech-Sabat G, et al. Preclinical evaluation of a COVID-19 vaccine candidate based on a recombinant RBD fusion heterodimer of SARS-CoV-2. iScience 2023: 106126.

24. Moros A, Prenafeta A, Barreiro A, et al. Immunogenicity and safety in pigs of PHH-1V, a SARS-CoV-2 RBD fusion heterodimer vaccine candidate. bioRxiv 2023: 2023.01.19.524684.

25. Prenafeta A, Bech-Sàbat G, Moros A, et al. Preclinical efficacy, safety, and immunogenicity of PHH-1V, a second-generation COVID-19 vaccine, in non-human primates. bioRxiv 2022: 2022.12.13.520255.

26. Wang Y, Liu C, Zhang C, et al. Structural basis for SARS-CoV-2 Delta variant recognition of ACE2 receptor and broadly neutralizing antibodies. Nat Commun 2022; 13(1): 871.

27. Chen J, Wang R, Gilby NB, Wei GW. Omicron (B.1.1.529): Infectivity, vaccine breakthrough, and antibody resistance. ArXiv 2021.

28. Leal L, Pich J, Ferrer L, et al. Safety and Immunogenicity of a Recombinant Protein RBD Fusion Heterodimer Vaccine against SARS-CoV-2: preliminary results of a phase 1-2a dose-escalating, randomized, double-blind clinical trial. medRxiv 2022: 2022.08.09.22278560.

29. Pradenas E, Trinité B, Urrea V, et al. Stable neutralizing antibody levels 6 months after mild and severe COVID-19 episodes. Med (N Y) 2021; 2(3): 313-20.e4.

30. Walsh EE, Frenck RW, Jr., Falsey AR, et al. Safety and Immunogenicity of Two RNA-Based Covid-19 Vaccine Candidates. N Engl J Med 2020; 383(25): 2439–50.

31. Lucaccioni H, Costa C, Duque MP, Balasegaram S, Sá Machado R. Risk of COVID-19 in Health Professionals: A Case-Control Study, Portugal. Portuguese Journal of Public Health 2021; 39(3): 137–44.

32. Cheng SMS, Mok CKP, Leung YWY, et al. Neutralizing antibodies against the SARS-CoV-2 Omicron variant BA.1 following homologous and heterologous CoronaVac or BNT162b2 vaccination. Nat Med 2022; 28(3): 486–9.

33. Graham BS. Rapid COVID-19 vaccine development. Science 2020; 368(6494): 945–6.

34. Hager KJ, Pérez Marc G, Gobeil P, et al. Efficacy and Safety of a Recombinant Plant-Based Adjuvanted Covid-19 Vaccine. N Engl J Med 2022; 386(22): 2084–96.

35. Sridhar S, Joaquin A, Bonaparte MI, et al. Safety and immunogenicity of an AS03-adjuvanted SARS-CoV-2 recombinant protein vaccine (CoV2 preS dTM) in healthy adults: interim findings from a phase 2, randomised, dose-finding, multicentre study. Lancet Infect Dis 2022; 22(5): 636–48.

36. Heath PT, Galiza EP, Baxter DN, et al. Safety and Efficacy of NVX-CoV2373 Covid-19 Vaccine. N Engl J Med 2021; 385(13): 1172–83.

37. Hielscher F, Schmidt T, Klemis V, et al. NVX-CoV2373-induced cellular and humoral immunity towards parental SARS-CoV-2 and VOCs compared to BNT162b2 and mRNA-1273-regimens. J Clin Virol 2022; 157: 105321.

