## Supplementary material for "Safety and immunogenicity of the protein-based PHH-1V compared to BNT162b2 as a heterologous SARS-CoV-2 booster vaccine in adults vaccinated against COVID-19: a multicentre, randomised, double-blind, non-inferiority phase IIb trial": Corominas et al 2022_supplementary

#### Supplementary methods

##### Randomization and masking

When an event may have been a Suspected Unexpected Serious Adverse Reaction (SUSAR), the blind should have been broken only for that specific subject. The blind should have been maintained for persons responsible for the ongoing conduct of the study (such as the management, monitors, Investigators) and those responsible for data analysis and interpretation of results at the conclusion of the study.

##### Withdrawal and Replacement of Subjects

A subject may discontinue study participation for any of the following reasons:

- If he/she is unwilling or unable to meet the protocol requirements.
- If the subject or the Investigator considers it best to end his/her participation in the study.
- Lost to follow-up.
- Withdrawal of consent.

All subjects have the right to withdraw their consent at any time during the study without prejudice to them. If possible, the subject withdrawing consent or discontinuing the study should complete an early termination visit. The date and reason for discontinuation or consent withdrawal will be documented in the eCRF. Subjects who withdraw or discontinue from the study will not be replaced. A subject will be considered lost to follow-up if he or she repeatedly fails to return for scheduled visits and is unable to be contacted by the study site. Reasonable efforts will be made, and documented, by site personnel to contact the subject to continue with their follow-up before determination that the subject is lost to follow-up.

##### Procedures

The vaccination diary was collected during the visit on day 14 and information collected about injection site reactions such as pain, tenderness, erythema/redness, induration/swelling, and systemic solicited event as fever, chills, nausea, malaise, vomiting, diarrhoea, headache, fatigue, muscle pain and joint pain.

The neutralisation titre against Wuhan-Hu-1 and the beta, gamma, delta and omicron variants were determined by inhibitory dilution 50 ($ID_{50}$) by a pseudovirion-based neutralisation assay (PBNA) and reported as reciprocal dilution for each individual sample and geometric mean titre (GMT) for treatment group comparison. The assay was performed at IrsiCaixa AIDS Research Institute (Badalona, Spain), using an HIV based Luciferase reporter pseudovirus pseudotyped with SARS-CoV-2 S protein.

Pseudoviruses were generated as described previously[1,2]. For the neutralisation assay, 200 $TCID_{50}$ of pseudovirus supernatant was preincubated with serial dilutions of the heat-inactivated serum samples for 45 minutes at 37°C and then added onto Human ACE2 overexpressing HEK293T cells. After 48 h, cells were lysed with britelite plus luciferase reagent (PerkinElmer, Waltham, MA, USA). Luminescence was measured for 0·2 s with an EnSight multimode plate reader (PerkinElmer). The neutralisation capacity of the serum samples was calculated by comparing the experimental RLUs calculated from infected cells treated with each serum to the max RLUs (maximal infectivity calculated from untreated infected cells) and min RLUs (minimal infectivity calculated from uninfected cells) and expressed as the neutralisation percentage:

Neutralisation (%) = (RLUmax–RLUexperimental)/(RLUmax–RLUmin) *100.

$ID_{50}$ were calculated by plotting and fitting neutralisation values and the log of serum dilution to a 4-parameters equation in Prism 9.0.2 (GraphPad Software, USA).

For the SARS-CoV-2 neutralisation test (VNA), SARS-CoV-2 were preincubated with serial 1/4 dilutions of heat-inactivated serum samples (ranging from 1/8 to a 1/8192 dilution) from the indicated individuals for 1 hour at 37 ºC. Pre-incubated viruses were added to 60000 Vero E6 cells per well in duplicate in 96 well plates. To control for serum-induced cytopathic effect, Vero E6 were also exposed to serial dilutions of the same serums but in the absence of virus. Seventy-two hours later, viral-induced or serum-induced cytopathic effect was measured using the Cell Titer Glo Luciferase reagent (Promega) and a Luminoskan Plate Reader (Thermo Fisher Scientific). The relative light units (RLU) were normalized to untreated noninfected cells (without serum or virus), and the ID50 (the reciprocal dilution inhibiting 50% of the cytopathic effect) was calculated by plotting and fitting the log of serum dilution vs. response to a 4-parameter equation in GraphPad Prism 9.3.1, as previously described in[1,3,4].

The cells, viruses, and viral titration were performed as follows; Vero E6 cells (ATCC CRL-1586) were cultured in Dulbecco's modified Eagle medium (Invitrogen) supplemented with 10% foetal bovine serum (FBS; Invitrogen), 100 U/ml penicillin, 100 μg/ml streptomycin (all from Invitrogen). Omicron or B.1.1.529 was isolated in Spain as described in (2). Genomic sequence was deposited at GISAID repository (http://gisaid.org) with accession ID EPI_ISL_8151031. Virus was sequenced as detailed in (2). Viral stocks were propagated in Vero E6 cells for two passages and titrated in 10-fold serial dilutions to calculate the Tissue Culture Infectious Dose ($TCID_{50}$) per mL. Infection was set to achieve a 50% of viral induced cytopathic effect measured with Cell Titer Glo Luciferase reagent, as described *below*.

The T-cell mediated immune responses against the SARS-CoV-2 Spike ("S") glycoprotein were assessed on cryopreserved PBMC at baseline and 2 weeks after receiving the boost by an IFN-γ ELISpot (IFN-γ ELISpot) and Intracellular Cytokine Staining (ICS). The cryopreserved PBMCs were thawed in RPMI complemented medium 20% FBS (R20) and then washed two times with RPMI 10% FBS (R10). Cells were counted and plated in a 96-wells round bottom plate using a total of $0{\cdot}5\times10^6$ cells per well. Next, PBMCs were stimulated with six peptide pools of overlapping SARS-CoV-2 peptides, each encompassing the SARS-CoV-2 regions S (2 pools) and RBD (4 pools covering Wuhan-Hu-1, alpha, beta, and delta variants), specified below:

- SPIKE_SA: 194 peptides overlapping the S1-2016 to S1-2196 region of the Spike protein from the ancestral Wuhan-Hu-1 strain.
- SPIKE_SB: 168 peptides overlapping the S1-2197 to S2-2377 region of the Spike protein from the ancestral Wuhan-Hu-1 strain.
- RBD: 84 peptides overlapping the RBD region of the Spike protein (Wuhan-Hu-1 sequence).
- RBD_B.1.1.7: 84 peptides overlapping the RBD region of the SARS-CoV-2 alpha variant.
- RBD_B.1.351: 84 peptides overlapping the RBD region of the SARS-CoV-2 beta variant.
- RBD_B.1617.2: 84 peptides overlapping the RBD region of the SARS-CoV-2 delta variant (this one only applies to ELISpot).

The PBMCs were incubated at a final concentration of 2·5 μg/mL per individual peptide pool. CEF peptide pool (composed of 23 peptides, which are MHC class I-restricted T-cell epitopes from human **C**ytomegalovirus, **E**pstein Barr virus and In**f**luenza virus -**CEF**- in the concentration 2·0 μg/ml, Mabtech, DK) was also used as positive control.

After overnight incubation, each well was washed 6 times with PBS and spot detection was accomplished by a two-step (biotinylated antibody/streptavidin-enzyme) antibody binding process; a 1-hour room temperature incubation with biotin plus anti-human IFN-γ, wash 6 times with PBS followed by another 1-hour incubation at room temperature with streptavidin. The wells were then incubated with developing solution, followed by 10 minutes at room temperature with 0·05% Tween 20 in PBS 1X and 6 washes with tap water. After drying upside down, ELISpots were read in the CTL reader system.

In parallel to the spot forming analysis, intracellular staining (ICS) was also performed with PBMCs incubated with different peptide pools. Hence, the PBMCs were incubated in the presence of 2 μg/mL of monoclonal antibodies against human CD28 (BD Pharmingen) and CD49d (BD Pharmingen) for 6 hours. During the last 4 hours of incubation, GolgiPlug (Brefeldin A, BD) was added to block cytokine transport. After incubation, PBMCs were washed with PBS 1X + 0·5% BSA + 0·1% sodium azide and incubated for 20 minutes with FcR Blocking Reagent (Milteny Biotec), then washed and stained for 25 minutes with the Live/Dead probe (LIVE/DEAD fixable near IR, Thermo Fisher Scientific) to discriminate dead cells as well as with surface antigens using the following antibodies: CD3 (PerCP), CD4 (BV421), CD8 (BV510) (BD Biosciences). Afterward, cells were washed twice in PBS 1X + 0·5% BSA + 0·1% sodium azide, fixed and permeabilized with Fix/Perm kit (BD) for intracellular cytokine staining. Cells were incubated again for 25 minutes with FcR Blocking Reagent (Milteny Biotec), washed and stained with anti-human antibodies of IFN-γ (APC), IL-2 (PE) and IL-4 (PECy7) (BD Biosciences). Finally, stained cells were washed twice with Perm/Wash 1X and fixed in formaldehyde 1%. Cytokine responses were background subtracted. All samples were acquired on BD FACSCanto II (BD Biosciences) flow cytometer and analysed using FlowJoTM v.10 software (Tree Star, Ashland, OR).

T-cell responses analysed by ELISpot were reported as the mean value of spot forming cells per $10^6$ PBMC (SFC/$10^6$ PBMC) upon stimulation with each peptide pool, after subtraction of background. In addition, intracellular cytokine staining (ICS) based T-cell assay was determined at different timepoints (baseline and 2 weeks after boost). ICS assays will include Th1/Th2 pathways (e.g., IL-2, IL-4, and IFN-γ) $CD4^+$ and $CD8^+$ T cell determinations using flow cytometry.

The safety assessment included the incidence and description of solicited local and systemic reactions, unsolicited local and systemic adverse events, serious adverse events, and adverse events of special interest. This is an ongoing study, and both solicited and unsolicited local and systemic adverse events were assessed through days 7 and 28, respectively, and safety laboratory parameters together with medically attended adverse events through the end of the study. In addition, severe infection was established and confirmed based on i) respiratory rate ≥30 breaths per minute, heart rate ≥125 beats per minute, oxygen saturation ($SpO_2$) ≤93% on room air at sea level or partial pressure of oxygen/fraction of inspired oxygen ($PaO_2/FIO_2$) <300 mm Hg, or ii) respiratory failure or acute respiratory distress syndrome (ARDS) defined as needing high-flow oxygen, non-invasive or mechanical ventilation, evidence of shock (systolic blood pressure <90 mmHg, diastolic blood pressure <60 mmHg that persist despite treatment with intravenous fluids or requiring vasopressors), or iii) significant acute renal, hepatic, or neurologic dysfunction, or iv) admission to an intensive care unit, or v) death.

**Statistical analysis**

Baseline characteristics

For the comparison of basal continuous variables, unpaired samples T-tests were used assuming data normality and homoskedasticity. For comparison of dichotomous variables between groups, the odds ratios of the corresponding proportions were estimated and tested against the null hypothesis $H_0$: OR = 1 using Fisher's Exact tests. The respective 95% confidence intervals of the estimated odds ratios were also calculated. Finally, categorical variables with more than two classes were also used for comparison between the two vaccination groups; in this case, Fisher's Exact Tests of independence were employed. Significance level was set to 5% in all tests.

Immunogenicity evaluation

Treatment group estimates and differences for ≥ 4-fold change response were analysed using a generalised estimating equations model for repeated measures. A scatter plot of $\log_{10}$-transformed titres with mean estimates and 95% CIs error bars by treatment group for each visit were produced. The weighted least square (LS) mean odds ratios for each treatment group were presented with the associated 95% CIs. The treatment group difference in weighted LS Means odds ratio (BNT162b2 active control vs PHH-1V) was also presented with the corresponding 95% CI and p-value for ratio = 1. Summary statistics for the fold change on day 14 post-boosting are presented for the modified intent-to-treat population. The generalised estimating equations model for repeated measures included the following effects: fixed effects, as treatment group, age group, visit and treatment-by-visit interaction, and repeated measures structure, as visits within subject. The model assumed a binomial family with logit link and an exchangeable working correlation structure. Weights were applied to the model estimation to account for sample distributions across covariates.

All statistical tests were performed using a two-tailed 5% overall significance level, unless otherwise stated, using SAS (Version 9.4) or R (Version 4.0.5).

Safety analysis

For comparison of dichotomous variables between groups, the odds ratios of the corresponding proportions were estimated and tested against the null hypothesis $H_0$: OR = 1 using Fisher's Exact tests. The respective 95% confidence intervals of the estimated ORs were also calculated. In all tests, significance level has been set to 5%.

All adverse events were coded using the MedDRA Version 24.1 coding system and displayed in tables and data listings by system organ class (SOC) and preferred term (PT).

Analyses of adverse events were performed for those events that are considered treatment-emergent, where treatment-emergent was defined as any adverse event with onset on or after the administration of study treatment through the end of the study (day 364) or any event that was present at baseline but worsened in intensity or was subsequently considered drug related by the Investigator through the end of the study.

Adverse events were summarised by subject incidence rates; therefore, in any tabulation, a subject contributed only once to the count for a given adverse event (SOC and PT).

A treatment-emergent adverse event (TEAE) was defined as an adverse event that started on or after the date of administration of study treatment until 28 days thereafter. This is an ongoing study, where TEAEs are shown until day 98 after boosting. If adverse event dates were incomplete and it was not clear whether the adverse event was treatment-emergent, it was assumed to be treatment-emergent. A treatment related adverse event was defined as related to the administration. If the TEAE had a missing relationship it was assumed to be related to all study treatments for analysis purposes.

The number and percentage of subjects with any TEAEs, with any TEAE assessed by the Investigator as related to treatment (related, probably related, possibly related, unlikely related, not related) and pooled related and unrelated categories with any SAE were summarised by treatment group and overall. Treatment-emergent adverse events by intensity (mild, moderate, and severe) and TEAEs leading to death were also summarised. In these tabulations, each subject contributed only once (i.e., the most related occurrence or the most intense occurrence) to each of the incidence rates in the descriptive analysis, regardless of the number of episodes.

Solicited local reactions and systemic events, as well as unsolicited local and systemic reactogenicity adverse events, are presented by intensity and cumulatively across severity levels.

Adverse events from baseline through day 14 after dosing are summarised and presented by intensity and cumulatively across severity levels. In addition, adverse events were summarised by maximum intensity and causal relationship to study drug. A separate summary of adverse event of special interest (AESIs), including potentially immune-mediated medical conditions (PIMMCs) and medically attended adverse events (MAAEs) through the end of study are to be reported.

No formal hypothesis testing analysis of adverse event incidence rates were performed. All adverse events occurring on-study including data collected via the subject diary were listed in subject data listings.

By-subject listings were also provided for the following: subject deaths, serious adverse events, and adverse events leading to study withdrawal.

**General methods**

All data listings that contained an evaluation date contained a relative study day (Study Day). Pre-treatment and on-treatment study days were numbered relative to the day of the first dose of study treatment which was designated as day 0. The preceding day is day -1, the day before that is day -2, etc.

All output were incorporated into Microsoft Word or Excel files, or Adobe Acrobat PDF files, sorted and labelled according to ICH recommendations, and formatted to the appropriate page size(s).

Tabulations were produced for appropriate demographic, baseline, efficacy, and safety parameters. For categorical variables, summary tabulations of the number and percentage of subjects within each category (with a category for missing data) of the parameter were presented. For continuous variables, the number of subjects, mean, standard deviation (SD), median and interquartile range, minimum, and maximum values were presented, where appropriate. Summarisations were presented by treatment arm and overall. For the immunogenicity variables, the geometric mean and geometric standard deviations were presented, as appropriate.

In the case where a variable was recorded as “> x”, “≥ x”, “< x” or “≤ x”, then for analysis purposes a value of x was taken. Where a range of values was quoted the midpoint of the range was taken.

All descriptive analyses were performed using SAS statistical software Version 9.4, unless otherwise noted. Statistical analyses were performed either using SAS Version 9.4 or R Version 4.0.5. In R, the following packages were used for the analysis of the baseline characteristics, safety, and immunogenicity data (quantitative analysis of binding antibodies, VNA, ELISpot and ICS): exact2x2 (Version 1.6.6), lme4 (Version 1.1-29), lmerTest (Version 3.1-3), nlme (Version 3.1-152) and emmeans (Version 1.7.4-1).

**References (Supplementary methods)**

1. Pradenas E, Trinité B, Urrea V, et al. Stable neutralizing antibody levels 6 months after mild and severe COVID-19 episodes. Med (N Y) 2021; 2(3): 313-20.e4.

2. Pradenas E, Trinité B, Urrea V, et al. Clinical course impacts early kinetics, magnitude, and amplitude of SARS-CoV-2 neutralizing antibodies beyond 1 year after infection. Cell Rep Med. 2022 Jan 24;3(2):100523.

3. Trinité B, Pradenas E, Marfil S, et al. Previous SARS-CoV-2 Infection Increases B.1.1.7 Cross-Neutralization by Vaccinated Individuals. Viruses 2021; 13(6).

4. Trinité B, Tarrés-Freixas F, Rodon J, et al. SARS-CoV-2 infection elicits a rapid neutralizing antibody response that correlates with disease severity. Sci Rep 2021; 11(1): 2608.

#### Supplementary results

In the 7 days after the boost administration, 257 subjects reported headache, where 157 (30·6%) were in the PHH-1V group reporting a mean (SD) duration of 2·0 (1·63) days, and 100 (39·7%) in the BNT162b2 group, reporting a duration of 1·8 (1·19) days. Additionally, in the same period 32 subjects reported fever, with 9 of them (1·75%) belonging to the PHH-1V group and reporting a mean (SD) duration of 1·7 (1·00) days, and 23 (9·12%) belonging to the BNT162b2 group, reporting 1·5 (0·90) days of fever.

### Supplementary Figure 1

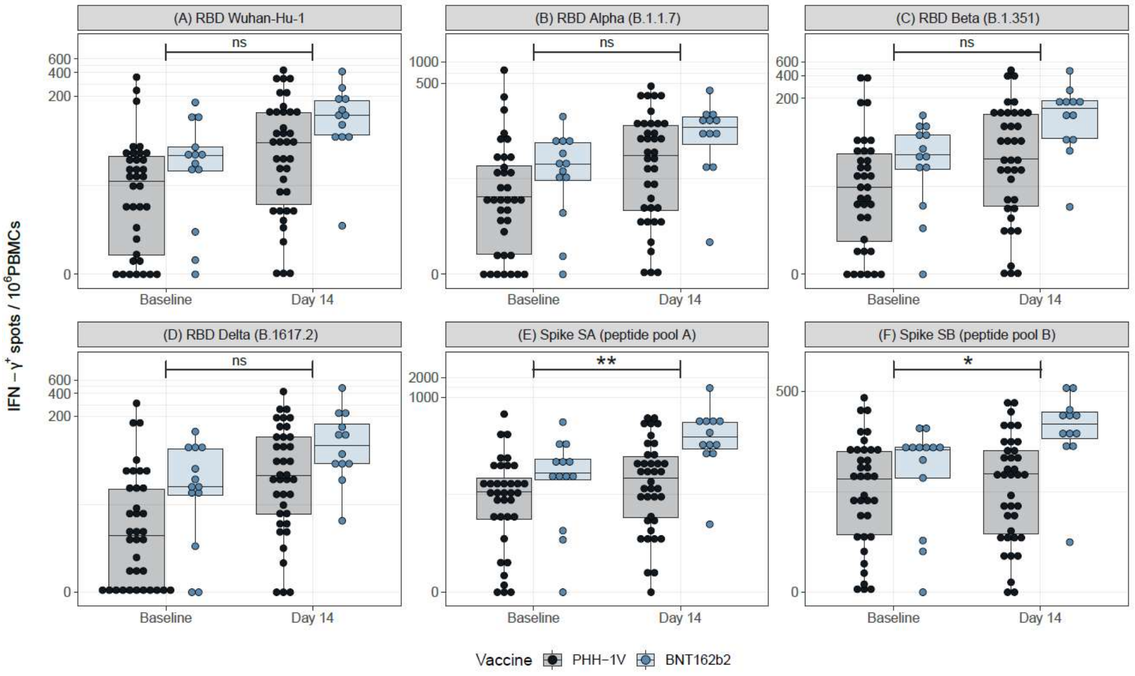

**Supplementary Figure 1:** Cellular SARS-CoV-2 specific immune response. PBMCs from participants receiving either PHH-1V (in grey) or BNT162b2 (in blue) were isolated before (Baseline) and two weeks after the boost immunization (Day 14). Results of IFN-γ ELISpot assay stimulating PBMCs with RBD and variants peptide pools [RBD (A); RBD B.1.1.7 (B); RBD B.1.351 (C) and RBD B.1.1617.2 (D)] and Spike [SA (E) and SB (F)] peptide pools are shown. Boxes depict the median (solid line) and the interquartile range (IQR), and whiskers expand each box edge 1.5 times the IQR. Interaction contrasts have been displayed in the plots, comparing the increase rates over time between the two vaccination groups. Non-significant differences in the increase rates between groups have been reported with "ns", while *p*-values lower than 0·05 indicate that the BNT162b2-vaccinated group has experienced a stronger boost compared to the PHH-1V arm.

*IQR=interquartile range; RDB; receptor binding domain for the SARS-CoV-2 spike protein (ancestor Wuhan-Hu-1 strain); RDB B.1.1.7 (Alpha variant); RDB B.1.351 (Beta variant); RDB B.1.1617.2 (Delta variant); Spike SA corresponds to 194 spike protein peptide pools overlapping the S1-2016 to S1-2196 region of the Spike protein; Spike SB corresponds to 168 spike protein peptide pools overlapping the S1-2197 to S2-2377 region of the Spike protein. Statistically significant differences are shown as * for p ≤ 0·05; ** for p ≤ 0·01. Non-significant comparisons have been indicated with "ns".*

#### Supplementary Table 1

Supplementary Table 1: Analysis of neutralizing and binding antibodies against SARS-CoV-2 variants on days 14, 28 and 98 post-vaccination boost in the mITT3(98) population.

| Variant | PHH-1V (n=410) | | | BNT162b2 (n=198) | | |
|---|---|---|---|---|---|---|
| | Day 14 | Day 28 | Day 98 | Day 14 | Day 28 | Day 98 |
| **Neutralizing antibodies** | | | | | | |
| **Wuhan-Hu-1** | | | | | | |
| n (%) | 407 (99·3) | 403 (98·3) | 78 (19·0) | 193 (97·5) | 195 (98·5) | 42 (21·2) |
| GMT | 2150·30 [1845·46, 2505·50] | 2393·53 [2053·79, 2789·48] | 1176·22 [924·71, 1496·14] | 3461·11 [2887·46, 4148·72] | 3058·96 [2553·30, 3664·76] | 986·47 [723·29, 1345·41] |
| GMT ratio | | | | 1·61 [1·35, 1·92]; p<0·0001 | 1·28 [1·07,1·52]; p=0·0062 | 0·84 [0·59, 1·20]; p=0·34 |
| GMFR | 23·31 [19·96, 27·23] | 25·95 [22·21, 30·32] | 12·75 [9·50, 17·11] | 41·06 [32·80, 51·41] | 36·29 [29·01, 45·40] | 11·70 [7·83, 17·48] |
| GMFR ratio | | | | 1·76 [1·43, 2·17]; p<0·0001 | 1·40 [1·14, 1·72]; p=0·0016 | 0·92 [0·63, 1·34]; p=0·65 |
| **Beta** | | | | | | |
| n (%) | 407 (99·3) | 403 (98·3) | 78 (19·0) | 193 (97·5) | 195 (98·5) | 42 (21·2) |
| GMT | 4738·87 [4072·39, 5514·43] | 4107·11 [3528·61, 4780·44] | 2015·79 [1572·75, 2583·65] | 2381·48 [2344·31, 3419·88] | 2574·09 [2132·42, 3107·23] | 1098·68 [792·71, 1522·76] |
| GMT ratio | | | | 0·60 [0·49, 0·73]; p<0·0001 | 0·63 [0·51, 0·77]; p<0·0001 | 0·55 [0·37, 0·80]; p=0·0022 |
| GMFR | 65·91 [56·07, 77·46] | 57·12 [48·57,67·17] | 28·03 [20·59, 38·17] | 46·07 [36·46, 58·20] | 41·88 [33·17, 52·87] | 17·87 [11·72, 27·25] |
| GMFR ratio | | | | 0·70 [0·56, 0·87]; p=0·0012 | 0·73 [0·59, 0·91]; p=0·0050 | 0·64 [0·43, 0·95]; p=0·0261 |
| **Delta** | | | | | | |
| n (%) | 407 (99·3) | 403 (98·3) | 78 (19·0) | 193 (97·5) | 195 (98·5) | 42 (21·2) |
| GMT | 1583·59 [1358·19, 1846·40] | 1835·50 [1573·92, 2140·55] | 1872·27 [1474·73, 2376·98] | 1525·92 [1270·12, 1833·23] | 1638·75 [1364·73, 1967·78] | 960·35 [705·13, 1307·67] |
| GMT ratio | | | | 0·96 [0·80, 1·16]; p=0·69 | 0·89 [0·74,1·07]; p=0·22 | 0·51 [0·36, 0·73]; p=0·0003 |
| GMFR | 33·11 [28·45, 38·53] | 38·38 [32·96, 44·69] | 39·15 [29·31, 52·28] | 37·17 [29·84, 46·30] | 39·92 [32·07, 49·69] | 23·39 [15·76, 34·72] |
| GMFR ratio | | | | 1·12 [0·92, 1·38]; p=0·26 | 1·04 [0·85, 1·27]; p=0·70 | 0·60 [0·41, 0·87]; p=0·0065 |
| **Omicron BA.1** | | | | | | |
| n (%) | 407 (99·3) | 403 (98·3) | 78 (19·0) | 193 (97·5) | 195 (98·5) | 42 (21·2) |
| GMT | 2283·10 [1929·00, 2702·21] | 1655·48 [1398·44, 1959·77] | 650·83 [503·86, 840·70] | 1331·54 [1091·36, 1624·56] | 998·46 [818·79,1217·55] | 357·43 [257·17, 496·78] |
| GMT ratio | | | | 0·58 [0·48, 0·71]; p<0·0001 | 0·60 [0·50, 0·73]; p<0·0001 | 0·55 [0·38, 0·80]; p=0·0020 |
| GMFR | 68·35 [58·2, 80·24] | 49·56 [42·19, 58·22] | 19·48 [14·35, 26·46] | 46·45 [36·82, 58·59] | 34·83 [27·63, 43·90] | 12·47 [8·21, 18·93] |
| GMFR ratio | | | | 0·68 [0·55, 0·84]; p<0·0004 | 0·70 [0·57, 0·87]; p=0·0013 | 0·64 [0·43, 0·95]; p=0·0257 |

*CI = confidence interval; GMT = Geometric Mean Titre; GMFR = Geometric Mean Fold Rise.*
*Data are shown for the mITT3(98) population, which includes all subjects in the mITT population without COVID-19 infections reported through Day 98.*

*n (%), refers to subjects with data; GMT is shown as adjusted treatment mean [95% CI]; GMT ratio is shown as BNT162b2 active control vs PHH-1V [95% CI] followed by p-value for ratio=1. GMFR is shown as fold rise of adjusted treatment means between timepoints [95% CI]; GMFR ratio is shown as BNT162b2 active control vs PHH-1V [95% CI] followed by p-value for ratio=1; GMFR fold change is shown for subjects with ≥4-fold change in binding antibodies; odds are shown as back-transformed adjusted treatment LS means [95% CI] ; Treatment effect is shown for "BNT162b2 vs PHH-1V" as the odds ratio [95% CI] followed by p-value for odds ratio=1.*

#### Supplementary Table 2

Supplementary Table 1: Solicited local reactions and systemic adverse events.

| | PHH-1V (N=513) | BNT162b2 (N=252) | OR (95% CI) | *p* value |
|---|---|---|---|---|
| **Solicited local reactions** | | | | |
| *Day 1* | | | | |
| Pain | 262 (51·1) | 176 (69·8) | 0·45 [0·32, 0·62] | 0·00 |
| Tenderness | 249 (48·5) | 160 (63·5) | 0·54 [0·39, 0·75] | 0·0001 |
| Induration/swelling | 30 (5·8) | 44 (17·5) | 0·29 [0·18, 0·48] | 0·00 |
| Erythema redness | 21 (4·1) | 25 (9·9) | 0·39 [0·21, 0·72] | 0·002 |
| *Day 3* | | | | |
| Pain | 42 (8·2) | 48 (19·0) | 0·38 [0·24, 0·60] | 0·00 |
| Tenderness | 42 (8·2) | 51 (20·2) | 0·35 [0·23, 0·55] | 0·00 |
| Induration/swelling | 10 (1·9) | 18 (7·1) | 0·26 [0·11, 0·60] | 0·0007 |
| Erythema redness | 10 (1·9) | 12 (4·8) | 0·4 [0·17, 0·96] | 0·04 |
| *Day 7* | | | | |
| Pain | 5 (1·0) | 4 (1·6) | 0·61 [0·16, 2·43] | 0·49 |
| Tenderness | 6 (1·2) | 5 (2·0) | 0·59 [0·17, 1·97] | 0·52 |
| Induration/swelling | 1 (0·2) | 2 (0·8) | 0·24 [0·01, 3·14] | 0·25 |
| Erythema redness | 2 (0·4) | 2 (0·8) | 0·49 [0·05, 4·55] | 0·60 |
| **Solicited Systemic Adverse Events** | | | | |
| *Day 1* | | | | |
| Fatigue | 82 (16·0) | 89 (35·3) | 0·35 [0·24, 0·50] | 0·00 |
| Headache | 73 (14·2) | 70 (27·8) | 0·43 [0·30, 0·63] | 0·00 |
| Muscle pain | 60 (11·7) | 74 (29·4) | 0·32 [0·22, 0·47] | 0·00 |
| Fever | 3 (0·6) | 18 (7·1) | 0·08 [0·02, 0·26] | 0·00 |
| Diarrhoea | 12 (2·3) | 4 (1·6) | 1·48 [0·48, 5·03] | 0·60 |
| Nausea | 8 (1·6) | 6 (2·4) | 0·65 [0·22, 1·90] | 0·41 |
| *Day 3* | | | | |
| Fatigue | 30 (5·8) | 12 (4·8) | 1·24 [0·61, 2·64] | 0·61 |
| Headache | 24 (4·7) | 13 (5·2) | 0·9 [0·45, 1·86] | 0·86 |
| Muscle pain | 15 (2·9) | 13 (5·2) | 0·55 [0·26, 1·26] | 0·15 |
| Fever | 2 (0·4) | 1 (0·4) | 0·98 [0·08, 28·59] | 1·00 |
| Diarrhoea | 5 (1·0) | 0 (0) | ∞ [0·49, ∞] | 0·18 |
| Nausea | 4 (0·8) | 1 (0·4) | 1·97 [0·26, 48·01] | 1.00 |
| *Day 7* | | | | |
| Fatigue | 10 (1·9) | 2 (0·8) | 2·48 [0·58, 15·91] | 0·35 |
| Headache | 19 (3·7) | 7 (2·8) | 1·35 [0·56, 3·44] | 0·67 |
| Muscle pain | 7 (1·4) | 1 (0·4) | 3·47 [0·49, 77·42] | 0·28 |
| Fever | 1 (0·2) | 0 (0) | ∞ [0.03, ∞] | 1·00 |
| Diarrhoea | 2 (0·4) | 0 (0) | ∞ [0.14, ∞] | 1.00 |

| | PHH-1V (N=513) | BNT162b2 (N=252) | OR (95% CI) | *p* value |
|---|---|---|---|---|
| Nausea | 1 (0·2) | 0 (0) | ∞ [0.03, ∞] | 1·00 |

*Data are shown for Day 1, 3 and 7 post booster vaccination, as "number of subjects (percentage)" in relation to the safety population. For comparison of dichotomous variables between groups, the OR of the corresponding proportions were estimated and tested against the null hypothesis $H_0$: OR = 1 using Fisher's Exact test. CI=Confidence Interval; OR=Odds ratio.*

#### Supplementary Table 3

Summary of Adverse Events by Treatment Group (Safety Population).

| | PHH-1V (N=513) | BNT162b2 (N=252) | OR (95% CI) | *p* value |
|---|---|---|---|---|
| **Total Adverse Events** | 1581 [458 (89·3)] | 1061 [238 (94·4)] | 0·49 [0·26, 0·91] | 0·0219 |
| Injection site pain | 748 [409 (79·7)] | 466 [225 (89·3)] | 0·47 [0·30, 0·75] | 0·0010 |
| Headache | 193 [160 (31·2)] | 122 [101 (40·1)] | 0·68 [0·49, 0·94] | 0·0190 |
| Fatigue | 166 [141 (27·5)] | 115 [106 (42·1)] | 0·52 [0·38, 0·72] | 0·0001 |
| Myalgia | 107 [100 (19·5)] | 93 [86 (34·1)] | 0·47 [0·33, 0·66] | 0 |
| Injection site induration | 45 [44 (8·6)] | 44 [43 (17·1)] | 0·46 [0·29, 0·72] | 0·001 |
| Injection site erythema | 33 [33 (6·4)] | 37 [36 (14·3)] | 0·41 [0·25, 0·70] | 0·0007 |
| **Intensity** | | | | |
| Mild | 1382 [342 (66·7)] | 885 [146 (57·9)] | 1·45 [1·06, 1.98] | 0·02 |
| Moderate | 187 [108 (21·1)] | 165 [85 (33·7)] | 0·52 [0·37, 0·74] | 0·0002 |
| Severe | 12 [8 (1·6) | 11 [7 (2·8)] | 0·55 [0·20, 1·74] | 0·27 |
| **Treatment-related Adverse Events** | 1384 [434 (84·6)] | 975 [231 (91·7)] | 0·5 [0·29, 0·83] | 0·0061 |
| **Serious Adverse Events (SAEs)** | 1 [1 (0·2)] | 0 [0 (0·0)] | ∞ [0·03, ∞] | 1 |
| **COVID-19 cases** | | | | |
| ≥ 14 days post-booster | 52 [52 (10·14)] | 31 [30 (11·9)] | 0·83 [0·51, 1·36] | 0·45 |

*Data are shown as the "total number of events [subjects (percentage)]" in relation to the safety population. For the total adverse events, is shown those events with a frequency ≥ 10% of treated patients, and as the system organ class preferred term. For comparison of dichotomous variables between groups, the OR of the corresponding proportions of affected individuals were estimated and tested against the null hypothesis $H_0$: OR = 1 using Fisher's Exact test. CI=Confidence Interval; OR=Odds ratio.*

#### Supplementary Table 4

Supplementary Table 3: Baseline characteristics the subset of participants (N=58) included in the analysis for neutralizing antibodies by infectious SARS-CoV-2 neutralisation test (VNA).

| | PHH-1V | BNT162b2 | Total | Confidence Interval | *p*-value |
|---|---|---|---|---|---|
| **Number of subjects, n (%)** | 34 (6.6) | 24 (9.5) | 58 (7.6) | | |
| **Age**, *years* | | | | | |
| All age groups, *median (range)* | 42.5 (22 – 64) | 54 (23 – 70) | 48 (22 – 70) | 9.04 [1.62, 16.46] | 0.0182 |
| 18-65, n (%) | 34 (100.0) | 20 (83.3) | 54 (93.1) | | |
| ≥ 65, n (%) | 0 (0.0) | 4 (16.7) | 4 (6.9) | ∞ (1.37, ∞) § | 0.0250 |
| **Sex** | | | | | |
| Female, n (%) | 23 (67.6) | 16 (66.7) | 39 (67.2) | | |
| Male, n (%) | 11 (32.4) | 8 (33.3) | 19 (32.8) | 1.04 [0.31, 3.20] § | 1.0000 |
| **Race** | | | | | |
| White, n (%) | 34 (100.0) | 24 (100.0) | 58 (100.0) | | |
| Asian, n (%) | 0 (0.0) | 0 (0.0) | 0 (0.0) | | |
| American Indian or Alaska native, n (%) | 0 (0.0) | 0 (0.0) | 0 (0.0) | | |
| Other, n (%) | 0 (0.0) | 0 (0.0) | 0 (0.0) | | 1.000 † |
| **Ethnicity** | | | | | |
| Hispanic or Latino, n (%) | 24 (70.6) | 16 (66.7) | 40 (69.0) | | |
| Not Hispanic or Latino, n (%) | 10 (29.4) | 8 (33.3) | 18 (31.0) | 1.20 [0.34, 3.84] § | 0.7799 |
| **BMI**, *median (range)* | 24.0 (18.8 – 39.8) | 24.2 (18.7 – 32.0) | 24.1 (18.7 – 39.8) | 0.87 [-1.28, 3.02] | 0.4219 |
| **Time between second dose and booster**, *median (range)* | 293 (189 – 337) | 291 (189 – 317) | 292 (189 – 337) | | 0.2882 ‡ |
| **Time boost to extraction, median (range)** | 14 (13 – 20) | 14 (13 – 15) | 14 (13 – 20) | | 0.2047 ‡ |

*Confidence intervals estimated using T-tests are shown as mean difference [95% confidence interval]. Where specified (§), confidence intervals estimated using Fisher's Exact test are shown as odds ratio [95% confidence interval].*

*† Data analysed by means of Fisher's Exact test.*

*‡ Data analysed by means of Anderson-Darling's test.*

#### Supplementary Information

Supplementary information – list of study centres

1. Hospital Germans Trias i Pujol
   Carretera de Canyet, s/n,
   08916 Badalona (Barcelona), Spain

   Principal investigator:
   Dr. José Moltó
  
   (+34) 934657602 Ext. 8887

2. Hospital Clínico Universitario de Valencia
   Av. de Blasco Ibáñez, 17,
   46010 València, Spain

   Principal investigator:
   Dr. Jorge Navarro
  
   (+34) 619207085

3. Hospital Regional Universitario de Málaga
   Av. de Carlos Haya, 84,
   29010 Málaga, Spain

   Principal investigator:
   Dra. Maria del Mar Vázquez
  
   (+34) 647771939

4. Hospital Principe de Asturias
   Carretera de Alcalá, s/n,
   28805 Meco (Madrid), Spain

   Principal investigator:
   Dr. Melchor Alvarez de Mons
  
   (+34) 918 87 81 00

5. Hospital de Cruces
   Cruces Plaza, s/n,
   48903 Barakaldo, Bizkaia, Spain

   Principal investigator:
   Dra. Eunate Arana
  
   (+34) 615709570

6. Hospital la Paz
   Paseo de la Castellana, 261,
   28046 Madrid, Spain

   Principal investigator:
   Dr. José Ramón Arribas
  

(+34) 918041950

7. Hospital General Universitario Gregorio Marañón
   Calle Dr. Esquerdo, 46,
   28007 Madrid, Spain

   Principal investigator:
   Dra. Patricia Muñoz
  
   (+34) 609227884

8. Hospital Josep Trueta
   Avinguda de França, S/N,
   17007 Girona, Spain

   Principal investigator:
   Dr. Rafael Ramos
  
   (+34) 607074712

9. Hospital Vall Hebron
   Passeig de la Vall d'Hebron, 119,
   08035 Barcelona, Spain

   Principal investigator:
   Dra. Susana Otero
  
   (+34) 669545766

10. Hospital Clínico de Barcelona
    C. de Villarroel, 170,
    08036 Barcelona, Spain

    Principal investigator:
    Dr. Alex Soriano
   
    (+34) 932275708
